# Genetic Determinants of Blood Cell Traits Influence Susceptibility to Childhood Acute Lymphoblastic Leukemia

**DOI:** 10.1101/2021.04.17.21255679

**Authors:** Linda Kachuri, Soyoung Jeon, Andrew T. DeWan, Catherine Metayer, Xiaomei Ma, John S. Witte, Charleston W. K. Chiang, Joseph L. Wiemels, Adam J. de Smith

**Affiliations:** Department of Epidemiology & Biostatistics, University of California San Francisco, San Francisco, CA; Center for Genetic Epidemiology, Department of Preventive Medicine, Keck School of Medicine, University of Southern California, Los Angeles, CA; Center for Perinatal, Pediatric and Environmental Epidemiology, Yale School of Public Health, New Haven, CT; Department of Chronic Disease Epidemiology, Yale School of Public Health, New Haven, CT; Division of Epidemiology & Biostatistics, School of Public Health, University of California Berkeley, Berkeley, CA; Institute for Human Genetics, University of California, San Francisco, San Francisco, USA; Department of Urology, University of California, San Francisco, San Francisco, USA; Department of Quantitative and Computational Biology, University of Southern California, Los Angeles, CA; Norris Comprehensive Cancer Center, University of Southern California, Los Angeles, CA

## Abstract

Acute lymphoblastic leukemia (ALL) is the most common childhood cancer. Despite overlap between genetic risk loci for ALL and hematologic traits, the etiological relevance of dysregulated blood cell homeostasis remains unclear. We investigated this question in a genome-wide association study (GWAS) of ALL (2666 cases, 60,272 controls) and multi-trait GWAS of 9 blood cell indices in the UK Biobank. We identified 3000 blood cell trait-associated (*P*<5.0×10^−8^) variants, explaining 4.0% to 23.9% of trait variation, and including 115 loci associated with blood cell ratios (LMR: lymphocyte/monocyte, NLR: neutrophil/lymphocyte, PLR: platelet/lymphocyte). ALL susceptibility was genetically correlated with lymphocyte counts (*r*_g_=0.088, p=4.0×10^−4^) and PLR (*r*_g_= −0.072, p=0.0017). In Mendelian randomization analyses, genetically predicted increase in lymphocyte counts was associated with increased ALL risk (Odds ratio (OR)=1.16, p=0.031) and strengthened after accounting for other cell types (OR=1.48, p=8.8×10^−4^). We observed positive associations with increasing LMR (OR=1.22, p=0.0017) and inverse effects for NLR (OR=0.67, p=3.1×10^−4^) and PLR (OR=0.80, p=0.002). Our study shows that a genetically induced shift towards higher lymphocyte counts, overall and in relation to monocytes, neutrophils, and platelets, confers an increased susceptibility to childhood ALL.

## INTRODUCTION

The hematopoietic system is remarkably orchestrated and responsible for some of the most important physiological functions, such as the production of adaptive and innate immunity, nutrient transport, clearance of toxins, and wound healing. Genetic factors contribute significantly to inter-individual variation in blood cell phenotypes, with heritability estimates for most blood cell traits ranging from 50-90% in twin studies to 30-40% in population-based studies of array-based heritability^1, 2, 3, 4^. Genome-wide association studies (GWAS) conducted in large population-based studies have revealed the highly polygenic nature of blood cell traits, with over 5,000 independently associated genetic loci identified to date^4, 5, 6^. These studies have provided insights into the genetic regulation of hematopoiesis and how dysregulation in blood cell development can lead to disease^7^. Genetic variants associated with blood cell variation have been implicated in the risk of immune-related conditions such as asthma, rheumatoid arthritis, and type 1 diabetes, and in rare blood disorders^4, 5, 6^. Positive genetic correlation was found between counts of varying blood cell types and the risk of myeloproliferative neoplasms, a group of diseases primarily of older age and characterized by the overproduction of mature myeloid cells^8^. However, the contribution of heritable variation in blood cell traits to the risk of other hematologic cancers has not been examined.

Acute lymphoblastic leukemia (ALL) is a malignancy of white blood cells, developing from immature B-cells or T-cells, and is the most common cancer diagnosed in children under 15 years of age^9^. Despite significant advances in treatment in recent decades and the corresponding improvements in survival rates^10^, ALL remains one of the leading causes of pediatric cancer mortality in the United States^11^. In addition, childhood ALL patients may endure severe toxicities during treatment, and survivors face long-term treatment-related morbidities and mortality^12, 13^. Thus, understanding the etiology of ALL remains important to identify avenues for disease prevention as well as potential novel treatment targets.

In most cases, the development of ALL is thought to follow a two-hit model of leukemogenesis, with *in utero* formation of a preleukemic clone and subsequent postnatal acquisition of secondary somatic mutations that drive progression to overt leukemia^14^. Epidemiologic studies have identified several genetic and non-genetic risk factors for ALL (reviewed in Williams et al.^15^ and Greaves^14^), but the biological mechanisms through which they promote leukemogenesis are largely unknown. GWAS of childhood ALL have revealed at least 12 common genetic risk loci to date, including at genes involved in hematopoiesis and early lymphoid development^16^, such as *ARID5B, IKZF1, CEBPE, GATA3, BMI1, IKZF3*, and *ERG* ^17, 18, 19, 20, 21, 22, 23, 24^. Intriguingly, several childhood ALL risk regions have also been associated with variation in blood cell traits^4, 6, 22, 23, 25^ and a recent phenome-wide association study (PheWAS) of childhood ALL identified platelet count as the most enriched trait among known ALL risk loci^26^. A comprehensive study of the role of blood cell trait variation in the etiology of childhood ALL is, therefore, warranted.

In this study, we utilize genome-wide data available from the UK Biobank (UKB) resource^27^ to perform a GWAS of blood cell traits and apply the discovered loci to a GWAS of childhood ALL in 2,666 cases and 60,272 controls of European ancestry. We assess the shared genetic architecture between blood cell phenotypes and childhood ALL and conduct Mendelian randomization (MR) and mediation analyses to disentangle putative causal effects of variation in blood cell homeostasis on ALL susceptibility.

## RESULTS

### Genetic determinants of blood cell traits

Genome-wide analyses revealed a substantial genetic contribution to blood cell trait variation. Heritability (*h*_g_) estimated from GWAS summary statistics on the full analytic cohort (median n=335,030) ranged from 3.1% for basophils to 21.8% for platelets (Figure 1, Supplementary Table 1). There was significant genetic correlation between all blood cell populations, which supports our rationale for using MTAG to leverage this shared genetic basis (Figure 2, Supplementary Table 2). Among non-composite traits, the largest correlations were observed between pairs of white blood cells: monocytes and neutrophils (*r*_g_=0.45, *P*=1.8×10^−83^), basophils and neutrophils (*r*_g_=0.44, *P*=4.0×10^−33^), and lymphocytes and monocytes (*r*_g_=0.41, *P*=1.3×10^−68^). Platelet counts were also significantly correlated with neutrophils (*r*_g_=0.24, *P*=1.7×10^−26^), lymphocytes (*r*_g_=0.22, *P*=3.8×10^−35^), and monocytes (*r*_g_=0.21, *P*=6.8×10^−26^). Cell type ratios LMR and NLR were primarily correlated with their component traits and with each other. PLR was significantly correlated with all phenotypes, including monocytes (*r*_g_=-0.20, *P*=2.5×10^−20^), neutrophils (*r*_g_=-0.16, *P*=2.7×10^−8^), basophils (*r*_g_=-0.21, *P*=1.1×10^−10^), and eosinophils (*r*_g_=-0.11, *P*=6.3×10^−7^).

**Figure 1:**
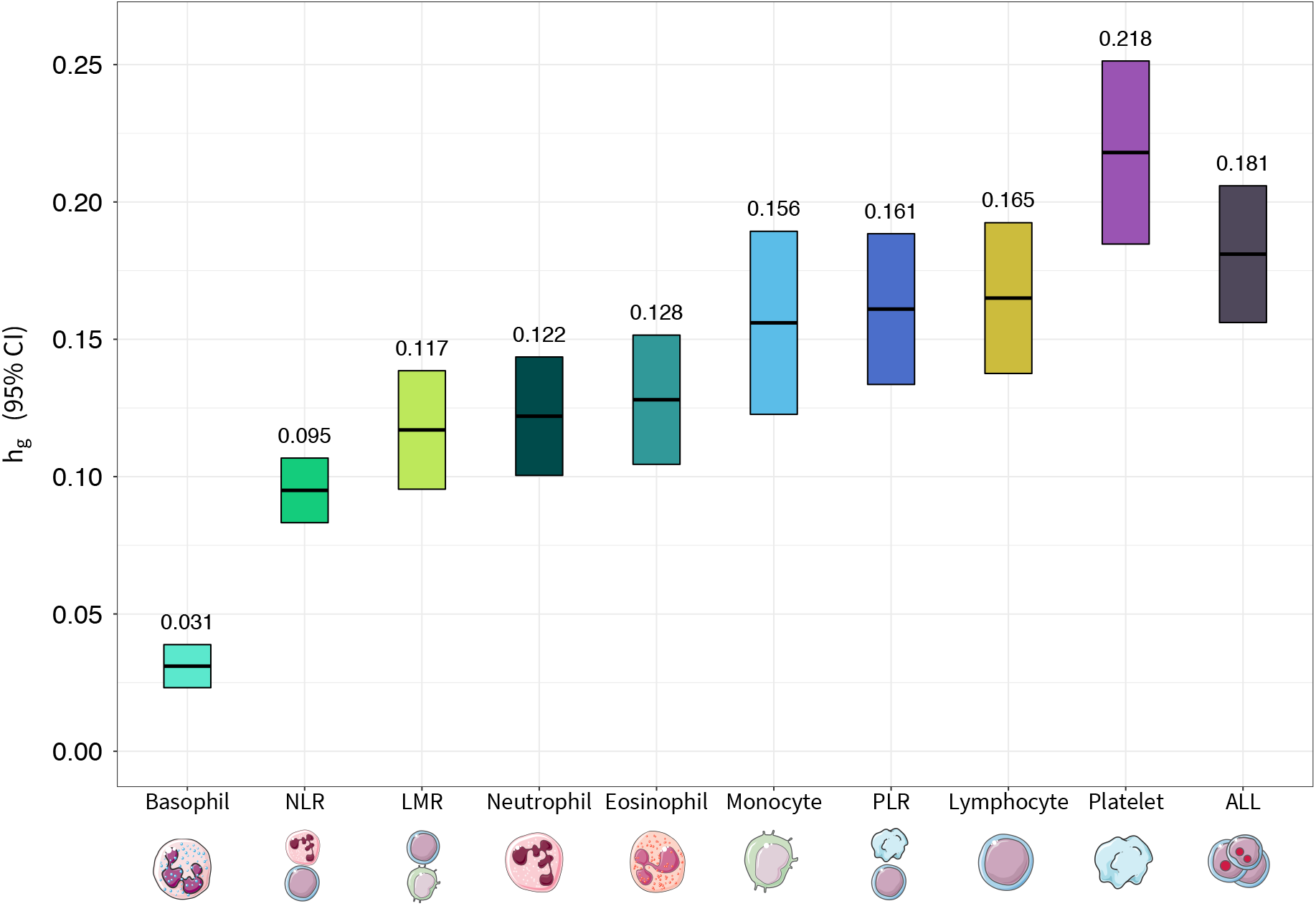
Array-based heritability (*h*_g_) estimates for acute lymphoblastic leukemia (ALL) and blood cell subtypes: lymphocytes, monocytes, neutrophils, basophils, eosinophils, basophils, platelets, lymphocyte to monocyte ratio (LMR), neutrophil to lymphocyte ratio (NLR), and platelet to lymphocyte ratio (PLR) estimated using LD score regression.

**Figure 2:**
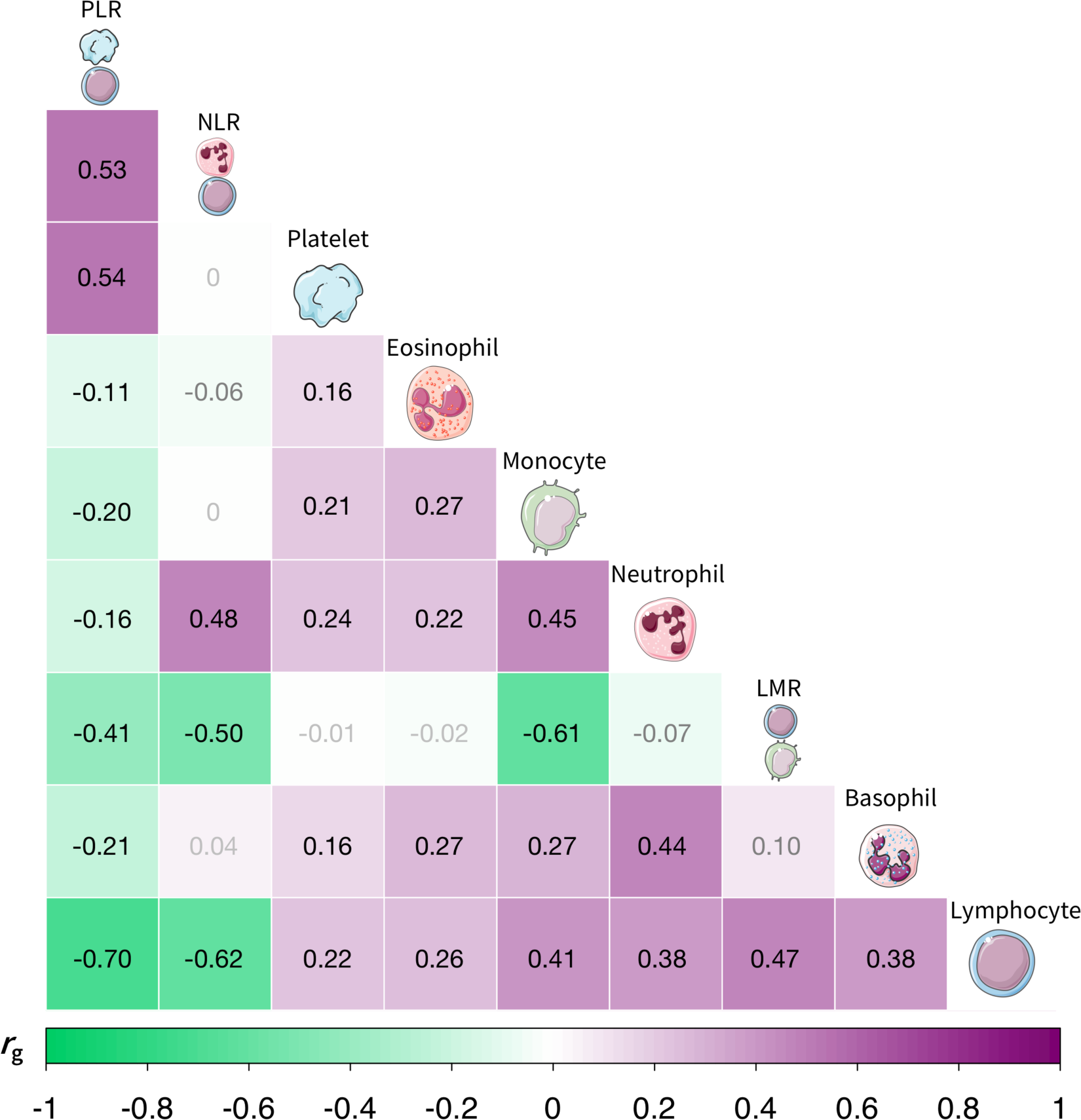
Genetic correlation (*r*_g_) estimates between blood cell subtypes: lymphocytes, monocytes, neutrophils, basophils, eosinophils, basophils, platelets, lymphocyte to monocyte ratio (LMR), neutrophil to lymphocyte ratio (NLR), and platelet to lymphocyte ratio (PLR) estimated using LD score regression. Associations with *P*<1.4×10^−3^ were considered statistically significant after Bonferroni correction for 36 pairs tested, with corresponding *r*_g_ estimates labeled in black font.

After applying our instrument selection criteria (discovery *P*_MTAG_<5×10^−8^, replication *P*_MTAG_<0.05; LD *r*^2^<0.05 within 10 Mb) we identified 3000 variants that were independent within, but not across hematological phenotypes (Supplementary Data 1). Of these, 2500 were associated with a single phenotype, 378 were associated with two, and 122 were instruments for 3 or more blood cell traits. The number of available instruments ranged from 157 for basophils to 692 for platelets (Supplementary Table 3). The proportion of trait variation accounted for by each set of instruments was estimated in the replication sample (100,284 to 100,764 individuals) and ranged between 5.1% for basophils to 24.4% for platelets (Supplementary Table 3). All traits had strong instruments with mean F statistics exceeding 60. Previous GWAS have not examined cell type ratios, while we identified 770 instruments specifically for ratio phenotypes: LMR, NLR, PLR. To assess whether these signals are captured by existing associations with cell counts or proportions, we performed clumping (LD *r*^2^<0.05 within 10 Mb) with loci reported in Vuckovic et al.^6^, a meta-analysis of UK Biobank and Blood Cell Consortium cohorts. This yielded 225 independent, ratio-specific variants in 115 cytoband loci, including six missense mutations (Figure 3, Supplementary Data 2).

**Figure 3:**
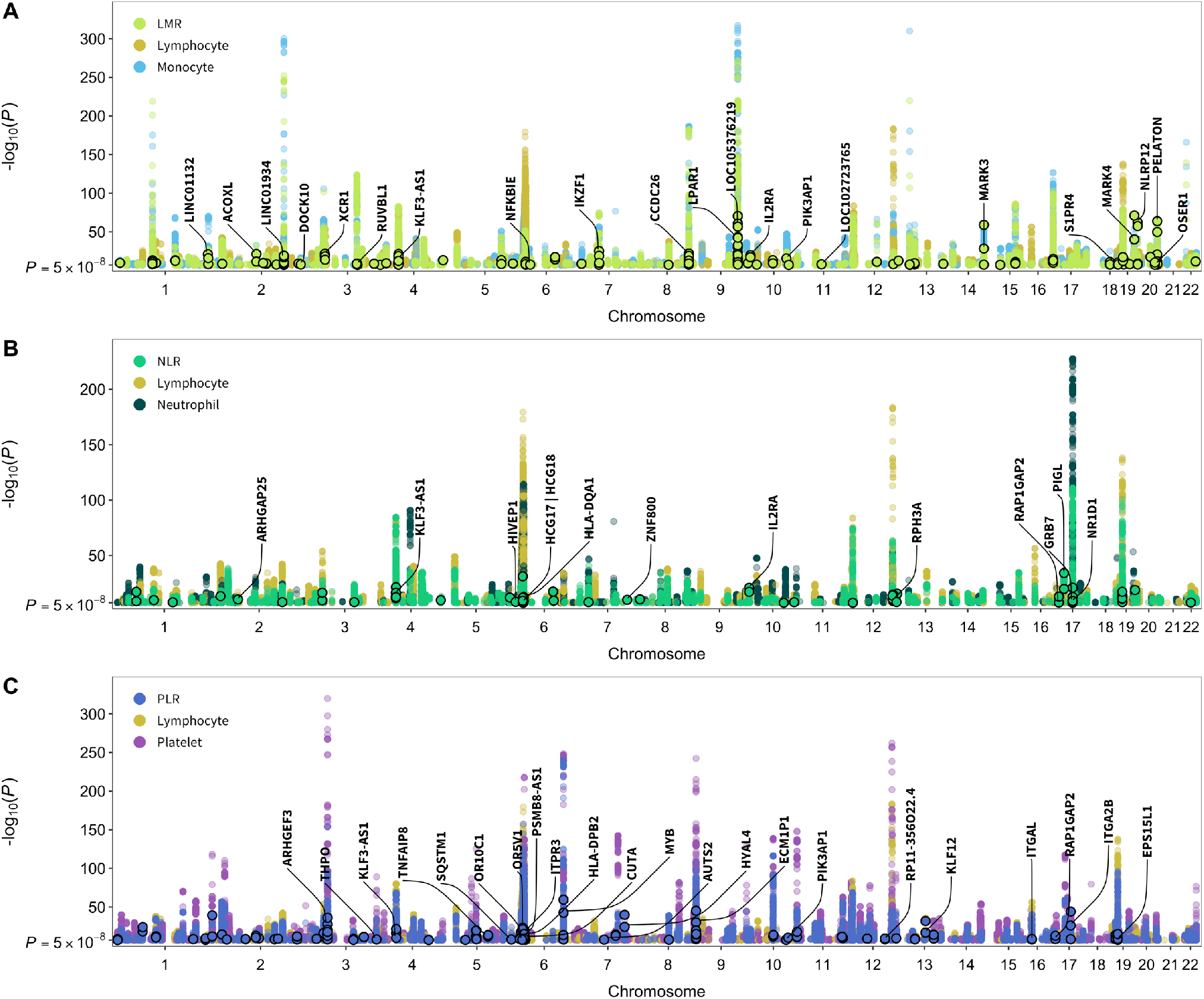
Genome-wide significant (*P*<5×10^−8^) associations for cell type ratios and their component count traits. Points with black borders denote variants that were selected only as instruments for the given ratio trait and are not in linkage disequilibrium (*r*^2^<0.05 within 10Mb) with previously reported loci for its component cell type counts or proportions. Labeled genes contain variants with specific functional features (CADD score >10, Regulome DB rank 1-3a, missense mutations) and/or *P*<1×10^−20^.

In-silico functional annotations identified overlap with multiple regulatory elements among all genetic instruments (Supplementary Data 1). A total of 324 variants were predicted to be in the top 10% of deleterious substitutions genome wide (CADD scores >10)^28^ and 138 had significant (p<0.05) evidence of overlap with open chromatin (FAIRE, DNAse, Pol-II, CTCF, and Myc) based on ENCODE data from up to 14 cell types. Over 80% of all instruments (n=2405) were expression quantitative trait loci (eQTL) in whole blood (FDR<0.05) based on results from eQTLGen^29^ (Supplementary Table 4). Fewer immune cell eQTLs were identified, although these reference datasets were much smaller. The highest proportion of eQTLs were observed in monocytes (27.0%), T-cells (23.6%), and neutrophils (21.1%), followed by B-cells (11.4%) (Supplementary Table 4). The proportion of immune cell eQTLs was broadly similar across categories of instruments, ranging from 26% for neutrophils to 16% for platelets (Supplementary Figure 2). For every instrument class, T-cell eQTLs were the most common. Lymphocytes were the most prevalent instrument class among cell-specific immune eQTLs.

Among instruments specific to one blood cell trait with eQTL effects in >2 cell types (Supplementary Figure 3A), the largest number of target genes was observed for platelet-specific and monocyte-specific instruments. Instruments for >4 blood cell traits with eQTL effects in >3 cell types (Supplementary Figure 3B) had a predominance of immune function genes in the HLA region and a previously identified ALL risk gene, *BAK1*. Among instruments for >5 blood cell traits with eQTL effects in a single cell/tissue type (Supplementary Figure 4) notable findings included multiple ALL risk genes (*IKZF3, CDKN2A, CDKN2B, IRF1*), and *FLT3*, a receptor tyrosine kinase that serves as a key regulator of hematopoiesis and is frequently mutated in ALL and acute myeloid leukemia (AML).

### Impact of blood cell variation on ALL risk

Associations between genetic determinants of blood cell trait variation and ALL susceptibility were investigated using a GWAS meta-analysis comprised of 2666 cases and 60,272 controls. Heritability of ALL was 18.1% (*h*_g_=0.181, SE=0.013), converted to the liability scale using SEER estimates of ALL lifetime risk in Non-Hispanic whites (0.15%) (Figure 1). At the genome-wide level, we observed positive correlations with ALL risk for increasing lymphocyte counts (*r*_g_=0.088, *P*=4.0×10^−4^), LMR (*r*_g_=0.065, *P*=0.012), and neutrophils (*r*_g_=0.051, *P*=0.027). Increasing PLR, corresponding to higher levels of platelets compared to lymphocytes, was inversely correlated (*r*_g_= −0.072, *P*=1.7×10^−3^) with ALL risk (Figure 4).

**Figure 4:**
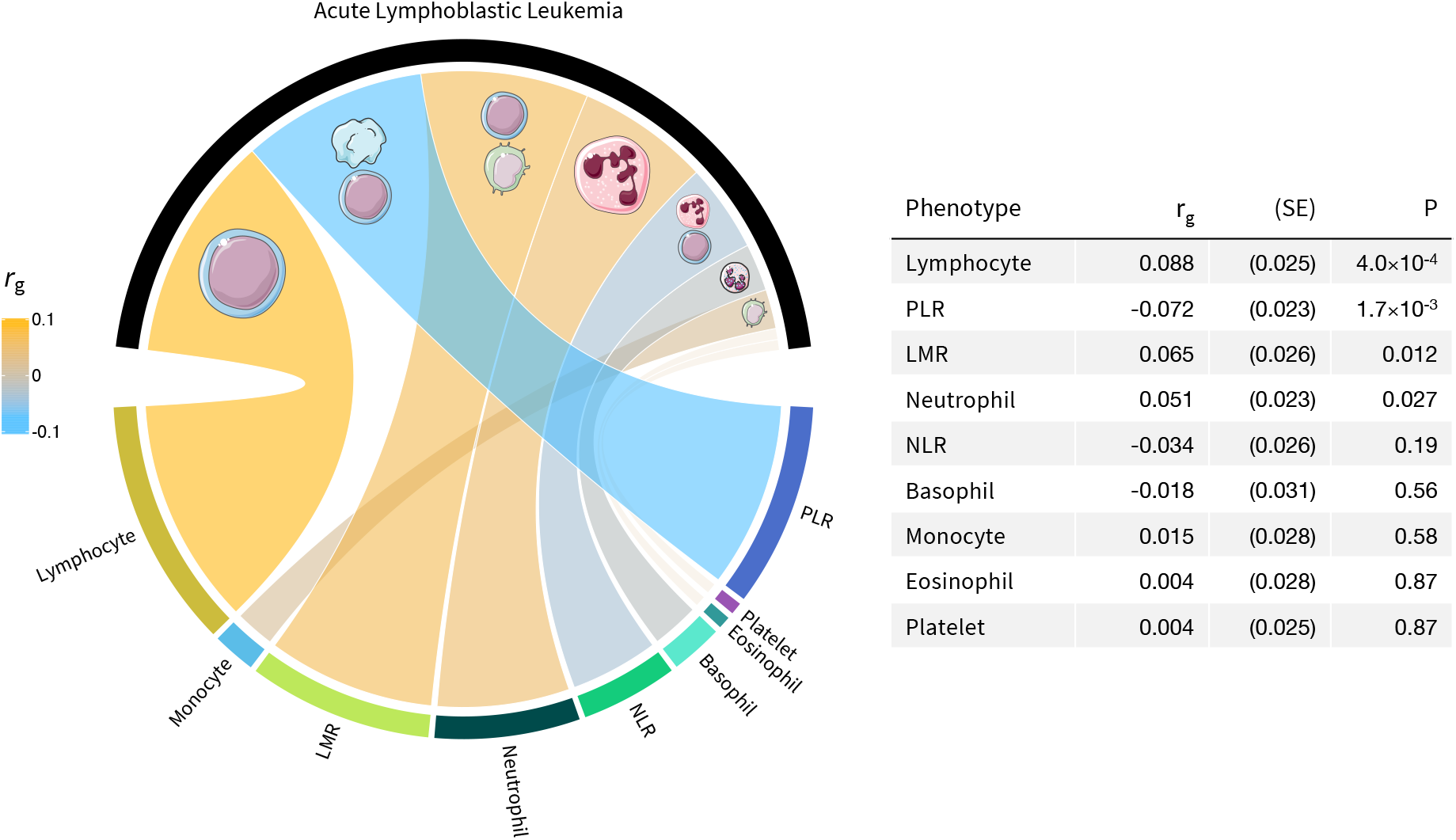
Genetic correlation (*r*_g_) between blood cell subtypes and acute lymphoblastic leukemia (ALL) based on genome-wide summary statistics. The colors in the circos plot correspond to the direction of genetic correlation, with warm shades depicting positive correlations between increasing blood cell counts or ratios and ALL risk, cool tones corresponding to inverse associations, and faded gray shades corresponding to null correlations. The width of each band in the circos plot is proportional to the magnitude of the absolute value of the *r*_g_ estimate.

Next, we conducted Mendelian Randomization (MR) analyses using genetic instruments developed in the UK Biobank to assess the putative causal relevance of blood cell trait variation in childhood ALL etiology (Figure 5, Supplementary Table 5). Primary analyses were based on the maximum likelihood (ML) and inverse variance weighted multiplicative random effects estimators (IVW-mre). To ensure that the observed results were robust against potential violations of MR assumptions, particularly horizontal pleiotropy, these estimators were complemented with shrinkage-based MR RAPS (Robust Adjusted Profile Score)^30, 31^ and MR pleiotropy residual sum and outlier (PRESSO) approaches^32^.

**Figure 5:**
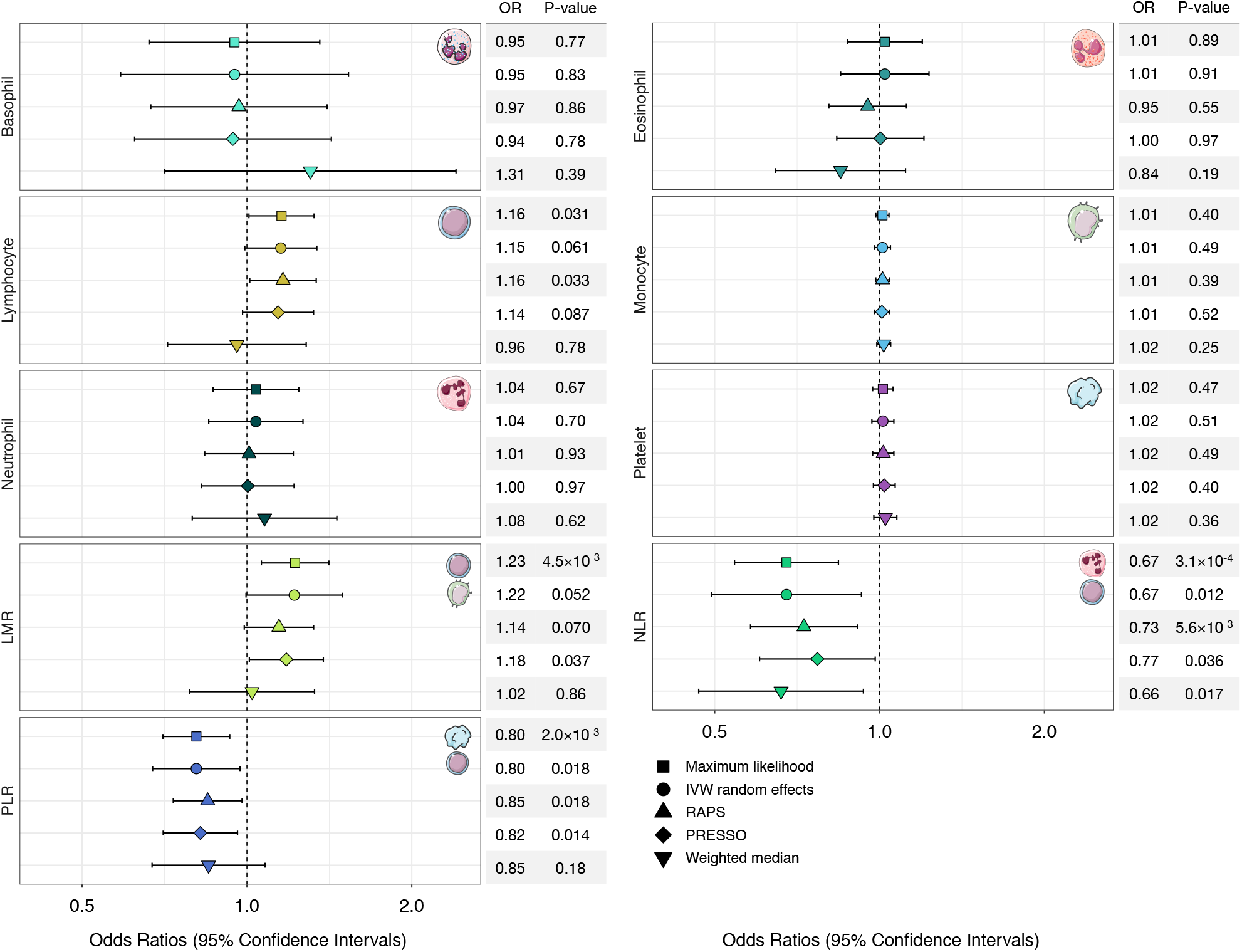
Odds ratios (OR) and 95% confidence intervals (CI) for the effect of increasing blood cell counts or cell type ratios on the risk of acute lymphoblastic leukemia (ALL), estimated using Mendelian Randomization (MR). Association results based on five different MR estimators are shown.

We did not detect evidence of directional horizontal pleiotropy based on the MR Egger intercept test for any blood cell traits (Supplementary Table 6). However, there was indication of balanced horizontal pleiotropy for all phenotypes based on Cochran’s Q (*P*_Q_<0.05) and the PRESSO Global test (*P*_Global_<0.05). Among white blood cells, a 1-SD increase in lymphocyte counts was associated with a modest increase in ALL risk (OR_ML_=1.16, 95% CI: 1.01-1.33, p=0.035; OR_IVW-mre_=1.15, 0.99-1.34, p=0.061). This effect was slightly attenuated in pleiotropy-corrected analyses (OR_PRESSO_=1.14, 0.98-1.32, p=0.087; OR_RAPS_=1.16, 1.01-1.34, p=0.033), but the effect size distortion was not significant (*P*_Dist_=0.88). There was no significant association between counts of other white blood cell types (monocytes, neutrophils, basophils, eosinophils) or platelets and ALL risk (Figure 4, Supplementary Table 5).

Considering ratios, indicating a genetic predisposition to a shift in the counts of one cell type relative to another, revealed several associations. An increase in LMR was associated with an approximately 22% increase in ALL risk (per 1-unit increase: OR_ML_=1.23, 1.07-1.41 p=4.5×10^−3^; OR_IVW-mre_=1.22, 1.00-1.50, p=0.052). Accounting for the influence of potentially pleiotropic outliers slightly attenuated this effect (OR_RAPS_=1.14, 0.99-1.32; OR_PRESSO_=1.18, 1.01-1.38). An inverse association with ALL risk was observed for increasing NLR (OR_ML_=0.67, 0.54-0.83, p=3.1×10^−4^; OR_IVW-mre_=0.67, 0.49-0.92, p=0.012), denoting a shift to higher levels of neutrophils compared to lymphocytes. Increased PLR was also associated with a lower risk of ALL (OR_ML_=0.80, 0.70-0.92, p=2.0×10^−3^; OR_IVW-mre_=0.80, 0.67-0.96, p=0.012). Associations with ALL for both phenotypes remained stable in sensitivity analyses correcting for pleiotropy (NLR: OR_PRESSO_=0.77, 0.66-0.98, p=0.036; PLR: OR_PRESSO_=0.82, 0.70-0.96, p=0.014) and outliers (NLR: OR_RAPS_=0.73, 0.58-0.91, p=5.6×10^−3^; PLR: OR_RAPS_=0.85, 0.73-0.98, p=0.025).

In addition to analytically correcting for pleiotropy, we also conducted analyses using a filtered set of genetic instruments excluding variants that showed evidence of heterogeneity based on Cochran’s Q in the observed causal effect estimates (Supplementary Table 7). These sensitivity analyses confirmed our previous findings showing that an increase in lymphocyte counts (OR_IVW-mre_=1.18, p=7.4×10^−3^) and LMR (OR_IVW-mre_=1.19, p=0.016) conferred a modest increase in ALL risk, while increased NLR (OR_IVW-mre_=0.67, p=7.4×10^−3^) and PLR (OR_IVW-mre_=0.82, p=5.8×10^−3^) were associated with lower risk.

Assessment of additional diagnostic tests indicated that our analysis was robust against main threats to validity (Supplementary Table 6). Associations were not affected by violations of the no measurement error assumption (*I*^2^_GX_>0.98). MR Steiger directionality test was used to orient the causal effects and confirmed that instruments for blood cell traits were affecting ALL susceptibility, not the reverse. The direction of this effect was robust across all traits, including lymphocytes (*P*=5.0×10^−135^), LMR (*P*=5.4×10^−98^), NLR (*P*=2.1×10^−10^), and PLR (*P*=1.2×10^−109^). Our analyses were powered to at least 80% to detect a minimum OR of 1.17 (equivalent to 0.85) for LMR and PLR, OR of 1.20 for lymphocytes, and OR of 1.28 (equivalent to 0.78) for NLR (Supplementary Figure 5).

Next, we conducted multivariable MR analyses to assess the degree to which specific blood cell subtypes exert independent direct effects on ALL (Supplementary Table 8). Among lymphocytes, monocytes, neutrophils, and platelets, only lymphocytes were independently associated with ALL (OR_MVMR_=1.18, 1.06-1.31, p=3.3×10^−3^) based on all variants and when restricting to instruments associated with each exposure (OR_MVMR-mod_=1.43, 1.16-1.76, p=8.8×10^−4^). This was confirmed using MV LASSO, which only retained lymphocytes. Among cell type ratios PLR was associated with ALL when considering all variants for all traits (OR_MVMR_=0.90, 0.82-0.99, p=0.033), but not in the instrument-specific analysis. PLR was the only trait selected by MV LASSO. Lastly, we explored the degree to which causal effects observed for ratio phenotypes were mediated by any of their component traits. We did not observe any statistically significant indirect effects, which suggests that the impact on ALL susceptibility observed for LMR, NLR, and PLR could not be attributed to effects on the counts of lymphocytes, monocytes, neutrophils, or platelets (Supplementary Table 9).

### Exploring mechanisms of ALL susceptibility

Lastly, MR-Clust^33^ was applied to blood cell traits associated with ALL to identify subgroups of variants with homogenous causal effects and novel ALL risk variants (Supplementary Figure 6; Supplementary Table 10). Clustering instruments for lymphocytes identified 10 variants indicating a large effect of increasing lymphocyte counts on ALL (OR=7.63). LMR instruments contained a cluster of 9 variants (OR=3.64). The largest cluster was identified for PLR, with 18 variants (OR of 0.27 per 1-unit increase in the ratio). A cluster comprised of 10 variants implied an extremely large inverse effect of NLR on ALL (OR=0.039). The substantive clusters were largely distinct, with one variant shared by all four traits (rs28447467: *P*_ALL_=0.026). Across all clusters, two variants were statistically significantly associated with ALL after correcting for the number of independent variants across all phenotypes tested (*P*_ALL_<5×10^−5^): rs6430608-C (OR=1.28, 1.15-1.41, *P*_ALL_=2.5×10^−6^) near *CXCR4* on 2q22.1 and rs76428106-C (OR=1.79, 1.36-2.35, *P*_ALL_=3.2×10^−5^) in *FLT3* on 13q12.2. The former is an intergenic variant specific to NLR with cis-effects on whole blood gene expression of *MCM6* (*P*_eQTL_=4.0×10^−28^) and *DARS1* (*P*_eQTL_=3.7×10^−37^), based on data from eQTLGen^29^. On the other hand, rs76428106, an intronic variant in *FLT3* and an eQTL for *FLT3* in whole blood (*P*_eQTL_=1.0×10^−11^)^29^ was included in substantive clusters for lymphocytes and PLR, but assigned to the “junk” cluster for LMR. Annotation of variants in substantive clusters revealed a predominance of previously reported associations with blood cell trait variation, as well as autoimmune and allergic conditions, such as type 1 diabetes, Crohn’s, asthma, and IgA deficiency (Supplementary Table 10).

Notable instruments assigned to “junk” clusters for LMR, PLR, and NLR included established ALL risk variants rs4948492 and rs4245597 (*ARID5B*, 10q21.2), rs2239630 (*CEBPE*, 14q11.2), rs78697948 (*IKZF1*, 7p12.2), and rs74756667 (8q24.2). These variants were also classified as outliers based on Cochran’s Q, suggesting that their effects on ALL susceptibility are predominantly mediated through pathways other than regulation of blood cell profiles. We formally tested this hypothesis using mediation analysis^34^ by decomposing the total SNP effect on ALL into direct and indirect (mediated) effects. For variants that were instruments for more than one blood cell phenotype, mediation was only explored for phenotypes significantly associated with ALL. Mediator-outcome effects were obtained from MR results excluding outliers (see Supplementary Table 7). For rs4245597 (*ARID5B*), only a small proportion of its effect on ALL risk was mediated by blood cell traits, 1.65% (0.79 – 2.51) was attributed to NLR and 0.84% (0.23 – 1.44) to PLR (Supplementary Table 11). Mediated effects attributed to LMR were not statistically significant for rs4245597 (*ARID5B*; 0.75%), rs2239630 (2.16%; *CEBPE*), and rs78697948 (1.72%; *IKZF1*).

Modest, but statistically significant, mediated effects were observed for putative novel ALL risk variants rs6430608 (NLR: 4.72%, 2.26 – 7.20) and rs76428106 (PLR: 2.43%, 0.68 – 4.19; lymphocytes: 2.51%, 0.67 – 4.35). The LMR-mediated effect of rs76428106 was larger (11.39%), but in the opposite direction from the effect of LMR on ALL, indicating pleiotropic effects consistent with the assignment of rs76428106 to the “junk” cluster for LMR. Of the six traits linked to this variant, its effect on monocytes (rs76428106-C: β=0.484, *P*=1.3×10^−310^) was by far the strongest. In MR analyses monocyte counts were not implicated in ALL susceptibility, suggesting that rs76428106 may be influencing ALL via other pathways or broad effects on hematopoiesis.

## DISCUSSION

Hematopoiesis is a tightly regulated hierarchical process designed to maintain optimal physiological ranges. Abnormalities in blood counts may be indicative of systemic inflammation or the presence of infections, and serve as indicators for a wide range of potentially adverse health conditions including inborn defects in hematopoiesis. While the responsive and sensitive nature of blood cell counts makes them useful clinical biomarkers, this poses a challenge for etiological studies. Elevated white blood cell counts are an established diagnostic feature of childhood ALL reflecting the overproduction of immature lymphocytes, or lymphoblasts, in the bone marrow. However, blood counts at a single time point, particularly in cancer cases, may not be representative of the individual’s stable, pre-diagnostic blood count profile, making it difficult to disentangle disease correlates from risk factors.

In this study, we leveraged the highly heritable nature of blood cell variation to evaluate its role in ALL pathogenesis without the limitations inherent in observational blood count measures. Our overarching finding is the convergence of genetic mechanisms resulting in increased lymphocyte counts and increased ALL susceptibility. Using genetic correlation and Mendelian randomization (MR) approaches, we observed a significant positive relationship between a genetically predicted increase in lymphocyte counts and ratio of lymphocytes to monocytes (LMR) and risk of ALL. Conversely, genetic predisposition to an increased ratio of platelets to lymphocytes (PLR) and neutrophils to lymphocytes (NLR) was inversely associated with ALL risk. These effects were largely robust to analytic corrections for horizontal pleiotropy and in some cases the removal of instruments contributing to heterogeneity strengthened the observed associations. Taken together, these results provide new insight into ALL etiology and point to a specific shift in blood cell homeostasis which confers an increased susceptibility.

However, the ways in which a genetic predisposition to over-production of lymphocytes may confer ALL risk are likely multifactorial. Stable and consistent causal effect estimates for lymphocytes, PLR, and NLR do not imply a single biological mechanism, even if they are estimated using valid instruments that primarily regulate the target blood cell trait. Acknowledging this, we propose two distinct, though not necessarily mutually exclusive, biological mechanisms related to the two-hit model of childhood ALL development that warrant further investigation. First, the initiating genetic lesions in childhood ALL, such as *ETV6*-*RUNX1* gene fusions, arise prenatally in most cases and require additional somatic mutations to progress to overt leukemia^14, 35^. The presence of common alleles across the spectrum of variants that subtly tune lymphocyte production may lead to an elevated risk of ALL by increasing the reservoir of preleukemic cell clones which, in turn, may increase the chances of acquiring “second-hit” oncogenic events and progression to ALL.

Second, the “delayed infection” hypothesis posits that children who lack early microbial exposures may have an unmodulated immune network that results in dysregulated immune responses to infectious stimuli later in childhood, and an increased risk of ALL^14^. This is supported by epidemiological evidence, with proxies for early-life infectious exposure, including daycare attendance and higher birth order, associated with a reduced risk of ALL^36, 37^, and by experimental models that demonstrated higher ALL incidence in mice with delayed exposure to pathogens^38, 39^. Further, children who develop ALL have been found to have different cytokine profiles at birth^40, 41^. Genetic variants that influence the blood cell phenotypes associated with ALL risk in our study may confer their effects via modulation of neonatal immune development and of immune responses to infections in childhood that may trigger ALL development. A shift towards increased lymphocytes to neutrophils is suggestive of increased adaptive immunity and lymphocyte activation in response to infections. This is consistent with our findings for NLR, a marker of increased inflammation, which was associated with reduced ALL risk. Similarly, reduced immune-inflammatory responses and increased activation of lymphocytes would be denoted by a higher ratio of lymphocytes to monocytes and lymphocytes to platelets^42^, both of which were associated with increased ALL risk in our study.

Previous studies have noted an overlap between ALL risk loci and genomic regions associated with blood cell phenotypes^21, 22, 23, 24^, however, ours is the first study to systematically analyze the contribution and causal effects of genetic variation across blood cell traits in ALL etiology. In a recent PheWAS, ALL risk variants were found to be enriched for regulation of platelet levels, but the overall association between platelet counts and ALL was null in Mendelian randomization and genetic score analyses using 223 platelet-associated variants^26^. This is consistent with our findings using over 600 genetic instruments for platelets, which indicate that variation in platelet counts alone does not influence ALL susceptibility, whereas PLR, which captures dysregulation of platelets in relation to lymphocytes, has a significant impact.

Indeed, we identified the cell type ratios LMR, NLR, and PLR as independent risk factors for ALL, with distinct genetic mechanisms not captured by their component traits. In multivariable MR analyses that concurrently modeled the effects of lymphocyte, monocyte, neutrophil, and platelet counts on ALL, lymphocytes remained as the only independent risk factor and this association with ALL strengthened compared to univariate analyses. However, there was no evidence that the total MR effects for LMR, NLR, and PLR were mediated either by lymphocytes or by the other cell populations. This implies that while dysregulation of lymphocyte homeostasis seems to be a key factor, it should be considered in the broader context of other blood cell subtypes.

In addition to identifying novel susceptibility pathways, our study also provides insights into the underlying mechanisms of several established ALL risk variants in *ARID5B, CEBPE, IKZF1*, and at chromosome 8q24. Despite significant associations with LMR and, in the case of *ARID5B*, with NLR and PLR, these variants were flagged as pleiotropic outliers in MR analyses, which was subsequently confirmed in mediation analyses. This supports that the overall effects of these loci on ALL risk are largely mediated by pathways other than regulation of blood cell trait variation, although we cannot rule out potential effects of these variants on early stages of hematopoiesis that may influence ALL development. Our MR clustering analysis also identified two putative novel ALL risk variants among genetic instruments for various blood cell traits: rs6430608 on 2q22.1 and rs76428106 in *FLT3* on 13q12.2.

While additional studies are needed to confirm their association with ALL, the locus at *FLT3* is of particular interest as this same variant was recently associated with an increased risk of autoimmune thyroid disease and AML^43^. The AML/ALL risk-increasing allele, rs76428106-C, has a frequency of approximately 1% in the general population (1.3% in UKB, 1.4% in ALL GWAS) and is reported as a splicing QTL in GTEx (*P*_sQTL_=1.3×10^−8^). Indeed, rs76428106-C was shown to generate a cryptic splice site resulting in truncation of the FLT3 protein, but an increase in FLT3 ligand levels^43^. Gain-of-function somatic mutations in *FLT3* are relatively frequent in childhood ALL^44^ and, while rs76428106 has greater effects on the production of myeloid cells than lymphocytes in our analyses, its putative effects on ALL risk may largely occur via activation of the RAS/MAPK pathway. This activation is likely to be restricted to key developmental decisions of hematopoietic cells given the delimited expression of FLT3 to hematopoietic stem and progenitor cells^45^. Less is known about the 2q22.1 variant, rs6430608, which is an eQTL for *MCM6* in blood and *CXCR4* in multiple tissues. *MCM6* is upregulated in multiple cancers and is believed to regulate DNA replication and activate MAPK/ERK signaling^46^. *CXCR4* is a chemokine receptor that facilitates HIV cell entry and regulates immune cell migration, including retention of B-cell precursors in the bone-marrow, and is being pursed as a therapeutic target in ALL and AML^47, 48^.

Several limitations of this study should be acknowledged. First, genetic instruments were developed for blood cell phenotypes measured in adult participants in the UK Biobank due to a paucity of adequately powered GWAS of blood cell traits in newborns or children. Environmental exposures throughout the life course influence blood cell dynamics, which has implications for the accuracy of genetic association estimates at different time points across the lifespan. While studies of blood cell development in pediatric populations should be pursued, the true underlying genetic architecture is not affected by age. This is also supported by studies of other complex traits, such as BMI, which showed that genetic risk scores developed in adults accurately predict weight gain in early childhood^49^. Therefore, we would expect any error in our genetic instruments developed in adults to bias MR results towards the null.

Second, our analysis was limited to broad classes of cell types, such as lymphocytes, and in future studies it will be important to distinguish between subpopulations of B-cell and T-cell lymphocytes. B-cell precursor ALL, the most common subtype, likely has distinct etiologic mechanisms from T-cell ALL^15^. Of relevance to our findings, the epidemiological evidence for the two-hit model of leukemogenesis is more compelling for B-cell ALL than for T-cell ALL^14^ and GWAS have revealed that hematopoietic transcription factor genes confer stronger effects on B-cell ALL risk^24^. We were also unable to characterize the effect of blood cell traits on B-cell ALL versus T-cell ALL or on specific molecular subtypes, or to explore the potential for germline-somatic interactions with specific ALL mutational signatures.

Finally, MTAG assumes that the variance-covariance matrix of effect sizes is homogeneous across all variants, which may be violated for SNPs that are null for one trait but non-null for other traits^50^. Replication is the best way to assess the credibility of observed associations, therefore our two-stage discovery and replication approach should minimize false positives. Furthermore, in a two-sample setting false positive instruments would bias Mendelian randomization estimates towards the null, not induce a spurious signal.

Despite some limitations, this study has important strengths that support the robustness of our findings. Our instrument development approach was optimized for Mendelian randomization studies of cancer etiology. The large sample size of the UK Biobank cohort allowed us to apply appropriate exclusions while retaining a sufficient number of participants for a two-stage discovery and replication analysis. Furthermore, applying the MTAG framework increased statistical power for identifying genetic determinants of specific blood cell traits, while taking into account the correlation between these phenotypes. This resulted in a set of strong genetic instruments explaining between 5% and 24% of variation in the target blood cell trait. These variants were enriched for multiple regulatory features, with over 80% having significant effects on gene expression in whole blood and up to 27% of instruments classified as eQTLs in immune cell subtypes, albeit with a limited degree of cell-type specificity in the eQTL effects across instrument classes. In addition, we characterized the genetic determinants of blood cell ratios, specifically LMR, NLR, and PLR, which have received considerably less attention in genetic association studies. A GWAS of PLR and NLR was conducted in 5901 healthy Dutch individuals, which identified one significant locus for PLR^51^. Examining these ratio phenotypes revealed novel ALL susceptibility pathways and helped contextualize the observed results for lymphocytes and platelets. Finally, the causal interpretation of our results depends on the validity of fundamental MR assumptions and, to this end, we employed a range of MR estimation methods with different underlying assumptions and conducted multiple diagnostic tests to interrogate the robustness of our results with respect to confounding, horizontal pleiotropy, and weak instrument bias.

In conclusion, we demonstrate for the first time that a genetic propensity for overproduction of lymphocytes, particularly in relation to other blood cell types, is associated with an increased risk of childhood ALL in Europeans. It will be important to elucidate the underlying biological mechanisms of our findings, and to assess their transferability to non-European populations.

## METHODS

### Population for Blood Trait Instrument Development

The UK Biobank (UKB) is a population-based prospective cohort of over 500,000 individuals aged 40-69 years at enrollment in 2006-2010 who completed extensive questionnaires on health-related factors, underwent physical assessments, and provided blood samples^27^. The UKB has been developed as a research resource and all participants provided informed consent at recruitment under the broad consent model. Four Beckman Coulter LH750 instruments were used to analyze blood samples collected in 4ml EDTA vacutainers. The LH750 instrument is a quantitative, automated hematology analyzer and leukocyte differential counter for in vitro diagnostic use in clinical laboratories. Samples were analyzed at the UK Biobank central laboratory within 24 hours of blood draw.

Quality control steps for this dataset have been previously described^52^. Briefly, genetic association analyses were restricted to individuals of predominantly European ancestry identified based on self-report and refined by excluding samples with any of the first two genetic ancestry principal components (PCs) outside of 5 standard deviations (SD) of the population mean. We removed samples with discordant self-reported and genetic sex, as well as one sample from each pair of first-degree relatives identified using KING ^53^. Using a subset of genotyped autosomal variants with minor allele frequency (MAF)≥0.01 and call rate ≥97%, we filtered samples with call rates <97% or heterozygosity >5 standard deviations (SD) from the mean, leaving 413,810 individuals available for analysis.

Additional exclusions were applied to optimize our dataset for developing genetic instruments for studies of cancer etiology by removing subjects with medical conditions that would alter blood cell proportions by pathophysiological conditions (n=13,597), such as pre-malignant myelodysplastic syndromes, autoimmune diseases, and immunodeficiencies, including HIV (Supplementary Figure 1). Blood counts (10^9^ cells/L) outside of the LH750 reportable range and extreme outliers (>99^th^ percentile) were excluded. Remaining values were converted to normalized z-scores with mean=0 and SD=1. In addition to overall blood cell counts we also examined relative concentrations: lymphocyte to monocyte ratio (LMR), neutrophil to lymphocyte ratio (NLR), and platelet to lymphocyte ratio (PLR).

### Genome-Wide Association Study of Blood Cell Traits

UKB participants were genotyped on the UK Biobank Affymetrix Axiom array (89%) or the UK BiLEVE array (11%) with imputation performed using the Haplotype Reference Consortium (HRC) and the merged UK10K and 1000 Genomes (1000G) phase 3 reference panels^27^. We excluded variants that were out of Hardy-Weinberg equilibrium in cancer-free individuals (*P*_HWE_<1×10^−5^) or had low imputation quality (INFO<0.30). Analyses were restricted to 10,369,434 variants with MAF≥0.005.

Genome-wide association analyses were conducted using linear regression in PLINK 2.0 (October 2017 version). Blood cell traits were analyzed using a two-stage GWAS, with a randomly sampled 70% of the cohort used for discovery and the remaining 30% reserved for replication, followed by Multi-Trait Analysis of GWAS (MTAG)^50^. Models for each trait were adjusted for age, age^2^, sex, genotyping array, the first 15 PC’s, cigarette pack-years, blood count device ID, and assay date. The resulting summary statistics were analyzed using MTAG, which has been shown to increase power to detect associations for correlated phenotypes ^50^. A key feature of this framework is distinguishing between different sources of correlation, such as genetic correlation and correlations due to sample overlap or biases in GWAS effect sizes due to population stratification or cryptic relatedness ^50^. MTAG integrates this information via a generalization of inverse-variance-weighted meta-analysis and outputs trait-specific effect estimates for each SNP ^50^. Genetic instruments were selected from MTAG results and defined as independent variants (linkage disequilibrium (LD) *r*^2^<0.05 in a clumping window of 10,000 kb) with *P*<5×10^−8^ in the discovery stage and *P*<0.05 and consistent direction of effect in the replication stage.

The functional relevance of the genetic instruments for blood cell traits was assessed using in-silico functional annotations: Combined Annotation Dependent Depletion (CADD) scores^28^ and Regulome DB^54^. We also explored associations with gene expression in whole blood in eQTLGen^29^, a meta-analysis of 31,684 subjects, and immune-cell specific effects in: DICE (Database of Immune Cell Expression)^55^, a dataset of 91 healthy blood donors; BLUEPRINT^56^ (n=197 healthy blood donors); and CEDAR (Correlated Expression and Disease Association Research)^57^ (n=322 healthy individuals from a cancer screening cohort). Gene expression datasets were accessed from the FUMA platform^58^: https://fuma.ctglab.nl.

### Childhood Acute Lymphoblastic Leukemia Datasets

Genetic associations with childhood ALL were obtained from a meta-analysis of 2666 cases and 60,272 controls from two separate genome-wide scans (details in Supplementary Note). The first GWAS consisted of a pooled dataset of 1162 cases and 1229 controls from the California Cancer Records Linkage Project (CCRLP)^21^ with 56,112 additional controls from the Kaiser Permanente Genetic Epidemiology Research on Ageing (GERA) cohort. Details of the CCRLP study and combined CCRLP/GERA GWAS have been previously described^21^, with the present analysis including additional GERA controls and imputation using the HRC reference panel (version r1.1 2016). In brief, newborn dried bloodspots were obtained from the California Biobank Program for CCRLP childhood ALL cases and cancer-free controls, identified via linkage between the statewide birth records maintained by the California Department of Public Health (1982-2009) and cancer diagnosis data from the California Cancer Registry (for the years 1988-2011). All CCRLP and GERA participants were genotyped on the Affymetrix Axiom World array. The second ALL GWAS included 1504 ALL cases from the Children’s Oncology Group (COG) and 2931 cancer-free controls from the Welcome Trust Case-Control Consortium (WTCCC), genotyped on either the Affymetrix Human SNP Array 6.0 (WTCCC, COG trials AALL0232 and P9904/9905)^25^ or the Affymetrix GeneChip Human Mapping 500K Array (COG P9906 and St. Jude Total Therapy XIIIB/XV)^59^. GWAS meta-analysis was restricted to individuals of predominantly European ancestry.

Standard quality control steps were implemented, removing variants with *P*_HWE_<1×10^−5^ in controls and imputation INFO<0.30. Additional filters were applied to minimize potential for bias due to the inclusion of external controls (Supplementary Figure 1). Variants associated with control group (CCRLP vs. GERA) at *P*<1×10^−5^ were removed (n=443). We also excluded variants if their MAF differed by >50% or ≥0.10 from the average MAF across CCRLP, GERA, and WTCCC controls (MAF≥0.05: n=3029; MAF<0.05: n=198,632). Lastly, allele frequencies in CCRLP/GERA and WTCCC controls were compared to the gnomAD non-Finnish European reference dataset and variants with absolute MAF differences ≥0.10 were filtered out (n=21,863).

### Heritability and Genetic Correlation

LD Score regression ^60^ was used to estimate heritability (*h*_g_) for each blood cell phenotype and for ALL, as well as the genetic correlation (*r*_g_) between each blood cell phenotype and ALL. We used a reference panel of LD scores generated from all variants that passed QC with MAF>0.0001 using a random sample of 10,000 European ancestry UKB participants. UKB LD scores were used to estimate *h*_g_ for each blood cell trait phenotype and *r*_g_ with ALL.

### Mendelian Randomization

Mendelian randomization (MR) analyses were carried out to investigate the potential causal relationship between blood cell trait variation and ALL. Genetic instruments excluded multi-allelic and non-inferable palindromic variants with intermediate allele frequencies (MAF>0.42). To minimize potential for bias due to differences in allele frequencies between exposure (UKB) and outcome (ALL) populations, analyses were restricted to variants with MAF≥0.01 and MAF difference<0.10. For instruments that were unavailable in the ALL dataset (n=294), LD proxies (*r*^2^>0.95) were obtained. MR analyses estimated odds ratios (OR) and corresponding 95% confidence intervals (CI) for a genetically predicted 1-SD increase in the normalized z-score for lymphocytes, monocytes, neutrophils, basophils, and eosinophils. For LMR, NLR, and PLR effects were estimated per 1-unit increase in the ratio.

Multiple MR estimators were used to strengthen inference by evaluating consistency in the observed effects. Maximum likelihood (ML) provides an unbiased estimate in the absence of horizontal pleiotropy, while inverse variance weighted multiplicative random-effects (IVW-mre) accounts for non-directional pleiotropy^61, 62^. Weighted median (WM)^63^ provides unbiased estimates when up to 50% of the weights are from invalid instruments. Shrinkage-based MR RAPS (Robust Adjusted Profile Score)^30, 31^ models additive pleiotropic effects and incorporates a robust loss function to limit the influence of invalid instruments. MR pleiotropy residual sum and outlier (PRESSO)^32^ regresses variant effects on the outcome on their exposure effects and compares the observed distance of all instruments to this regression line with the expected distance under the null hypothesis of no horizontal pleiotropy^32^.

To assess potential violations of MR assumptions we examined the following diagnostic tests: i) deviation of the MR Egger intercept from 0 (p<0.05) indicative of directional horizontal pleiotropy; ii) *I*^2^_GX_<0.90 indicative of regression dilution bias due to exposure measurement error which may inflate the MR Egger pleiotropy test^64^; iii) Cochran’s Q-statistic *P*_Q_<0.05 or MR PRESSO *P*_Global_<0.05 indicative of heterogeneity due to balanced horizontal pleiotropy. We also report MR PRESSO distortion p-values, which test for significant differences between the original and pleiotropy-corrected effect estimates.

Next, we conducted multivariable MR (MVMR) analyses to estimate direct effects of specific blood cell traits on ALL, after accounting for related phenotypes. MVMR requires SNP effects for all instruments across all exposures so that they can be regressed against the outcome together, weighting for the inverse variance of the outcome (MVMR-IVW). We also applied a modified analysis where the instruments are selected for each exposure based on *P*<5×10^−8^ and then all exposures for those SNPs are regressed together (MVMR-IVW_mod_). Feature selection was also performed using MV LASSO.

For ratios we conducted summary-based mediation analysis to decompose the observed total MR effects into direct and indirect effects mediated by each of the component traits^65^. For instance, for LMR we quantified indirect effects on ALL risk that were mediated through regulation of lymphocyte and monocyte counts, respectively, as well as direct LMR effects on ALL.

Lastly, we applied MR-Clust^33^, a heterogeneity-based clustering method for detecting distinct values of the causal effect that are evidenced by multiple genetic variants. MR-Clust assigns variants to *K* substantive clusters where all variants indicate the same causal effect, a null cluster, and a “junk” cluster, which includes non-null variants which do not fit into any of the substantive clusters. This approach may reveal different causal or pleiotropic pathways and identify potentially novel ALL risk variants due to a reduced burden of multiple testing compared with GWAS. Variants were assigned to a cluster if their conditional probability of cluster membership was greater than 0.50. Clusters were formed with a minimum of 4 variants.

All statistical analyses were conducted using R (version 4.0.2). Mendelian randomization analyses were conducted using the TwoSampleMR R package (version 0.5.5).

## Data Availability

This research was conducted with approved access to UK Biobank data under application number 14105 (PI: Witte) and in accordance with the UK Biobank Ethics and Governance Framework. UK Biobank data are publicly available by request from https://www.ukbiobank.ac.uk. Ethics approval for establishing the UK Biobank resource was obtained from the North West Centre for Research Ethics Committee (11/NW/0382).

This study included the analysis of data derived from biospecimens from the California Biobank Program (CCRLP study). Any uploading of genomic data and/or sharing of these biospecimens or individual data derived from these biospecimens has been determined to violate the statutory scheme of the California Health and Safety Code Sections 124980(j), 124991(b), (g), (h), and 103850 (a) and (d), which protect the confidential nature of biospecimens and individual data derived from biospecimens. This study was approved by Institutional Review Boards at the California Health and Human Services Agency, University of Southern California, Yale University, and the University of California San Francisco. The de-identified newborn dried blood spots for the CCRLP (California Biobank Program SIS request # 26) were obtained with a waiver of consent from the Committee for the Protection of Human Subjects of the State of California.

This study makes use of data from the Kaiser Permanente (KP) Research Program on Genes, Environment and Health (RPGEH) Genetic Epidemiology Research on Adult Health and Aging (GERA) cohort, available from dbGaP (Study Accession: phs000788.v1.p2).

This study also makes use of data generated by the Wellcome Trust Case-Control Consortium available by request from the European Genotype Archive: http://www.ebi.ac.uk/ega (Study Accession: EGAD00000000021). Genotype data for COG ALL cases are available for download from dbGaP (Study Accession: phs000638.v1.p1).

## ACKNOWLEDGEMENTS

This work was supported by research grants from the National Institutes of Health (NIH) National Cancer Institute (NCI): R03CA245998 (AJD and LK), K99CA246076 (LK), R01CA155461 (JLW, XM) and R01CA175737 (JLW, XM). The content of this manuscript is solely the responsibility of the authors and does not necessarily represent the official views of the National Institutes of Health.

The collection of cancer incidence data used in this study was supported by the California Department of Public Health pursuant to California Health and Safety Code Section 103885; Centers for Disease Control and Prevention’s (CDC) National Program of Cancer Registries, under cooperative agreement 5NU58DP003862-04/DP003862; the National Cancer Institute’s Surveillance, Epidemiology and End Results Program under contract HHSN261201000140C awarded to the Cancer Prevention Institute of California, contract HHSN261201000035C awarded to the University of Southern California, and contract HHSN261201000034C awarded to the Public Health Institute. The ideas and opinions expressed herein are those of the author(s) and do not necessarily reflect the opinions of the State of California, Department of Public Health, the National Cancer Institute, and the Centers for Disease Control and Prevention or their Contractors and Subcontractors. A subset of the CCRLP data used in this study were obtained from the California Biobank Program at the California Department of Public Health (CDPH), SIS request number 26, in accordance with Section 6555(b),17CCR. The CDPH is not responsible for the results or conclusions drawn by the authors of this publication.

---

**Supplementary Figure 1:**
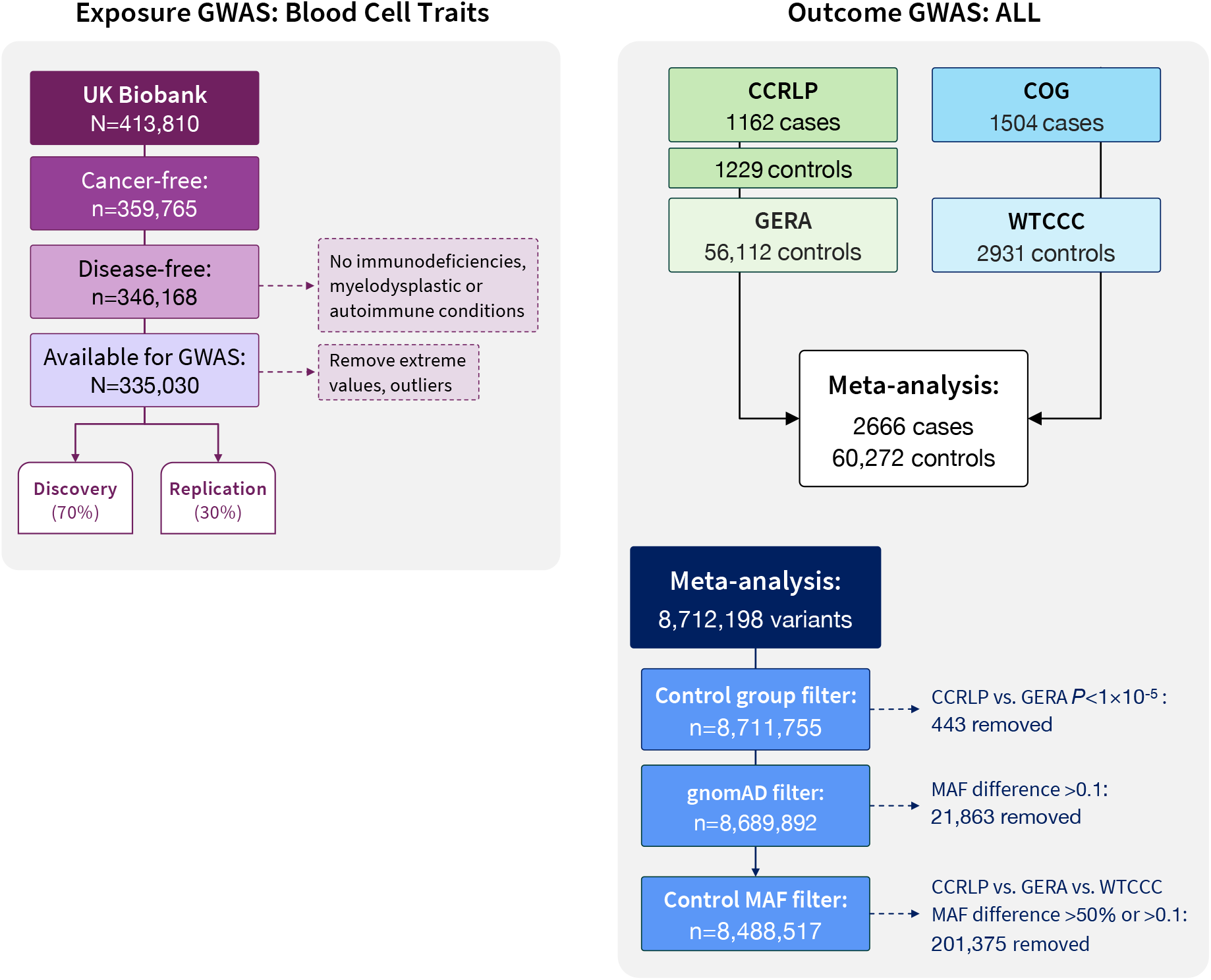
Flowchart outlining the study populations and additional quality control steps. Participants were restricted to individuals of predominantly European ancestry.

**Supplementary Figure 2:**
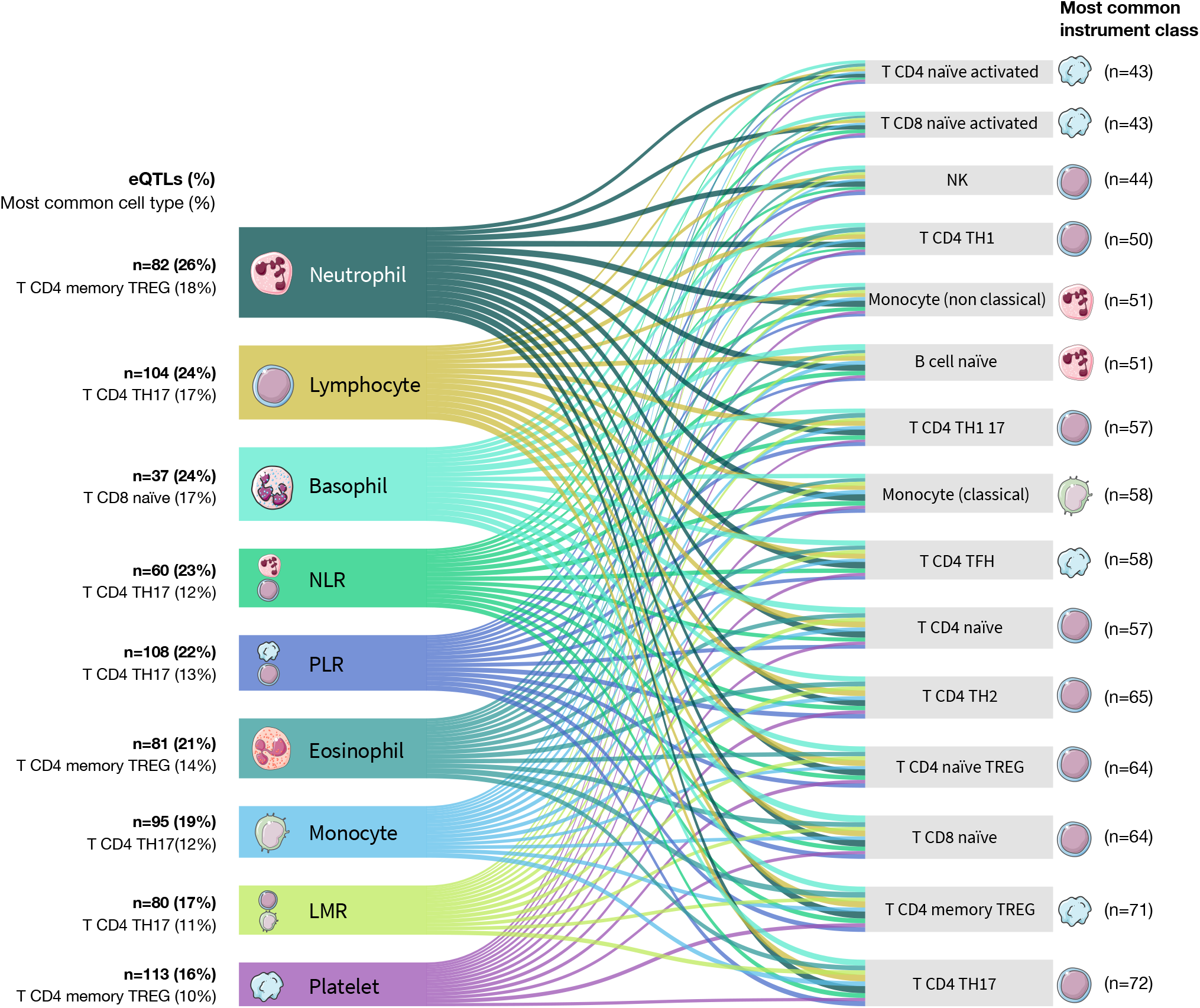
Sankey style plot illustrating the pattern of cell-type specific effects on gene expression obtained from DICE (Database of Immune Cell Expression) for blood cell trait genetic instruments. Genetic instruments were considered expression quantitative trait loci (eQTL) if their associations with gene expression were significant at FDR<0.05. The color of each band corresponds to the blood cell type and the width of each band is scaled to reflect the proportion of all eQTL signals accounted for by a specific cell type.

**Supplementary Figure 3:**
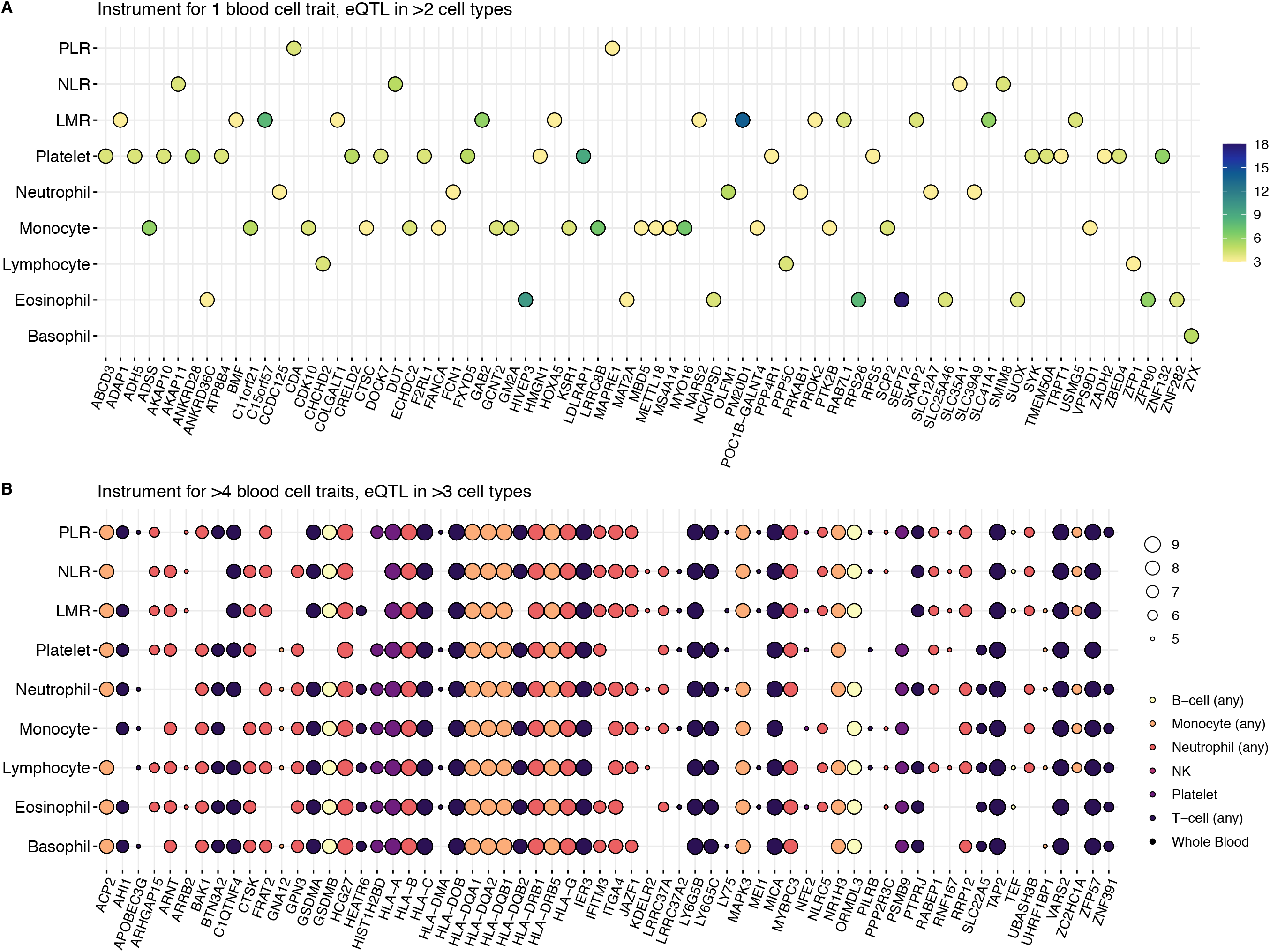
Genes with expression trait loci (eQTL) in multiple cell types found among instruments for blood cell traits

**Supplementary Figure 4:**
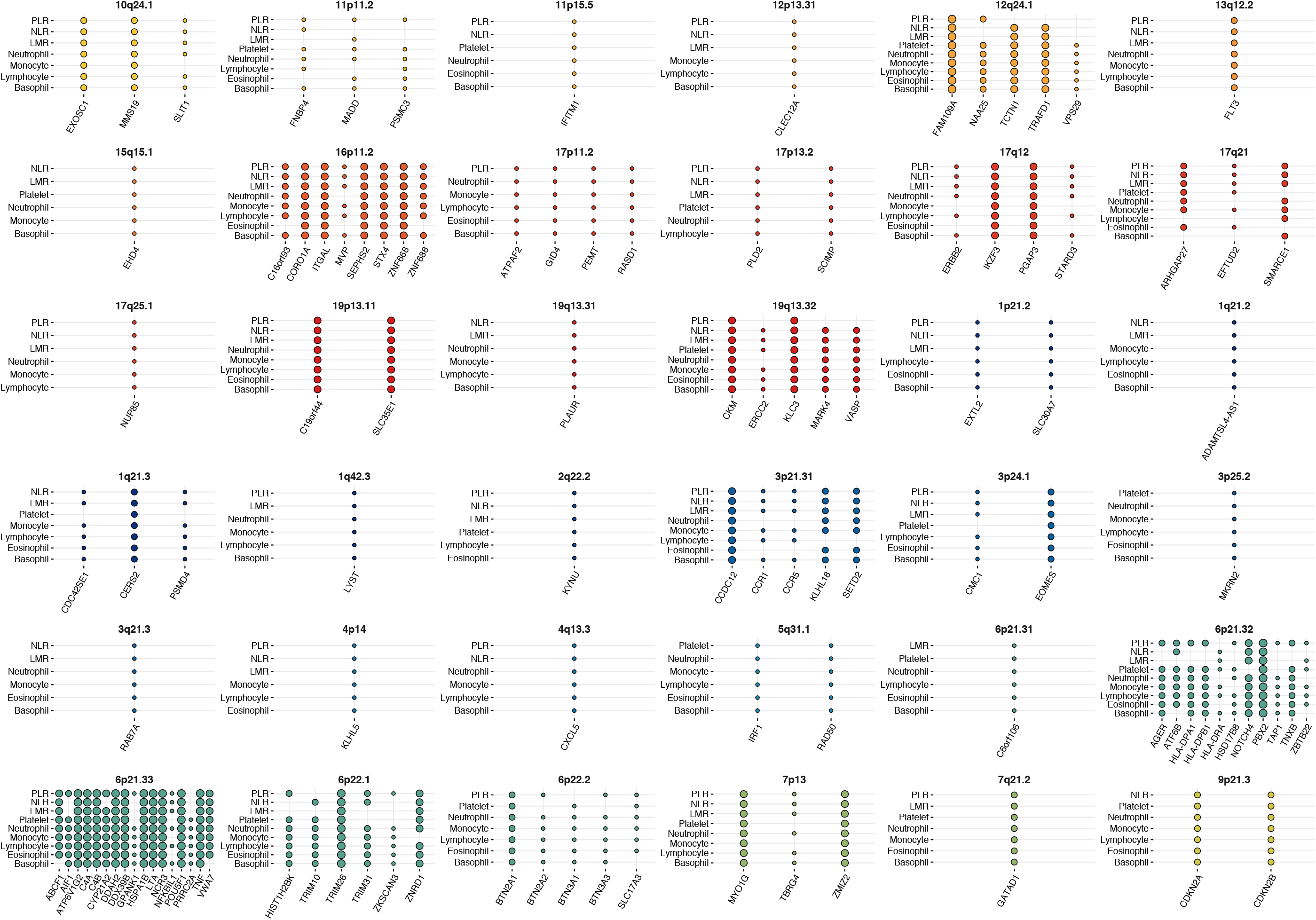
Genes with expression trait loci (eQTL) in one cell type found among genetic instruments for >5 blood cell traits

**Supplementary Figure 5:**
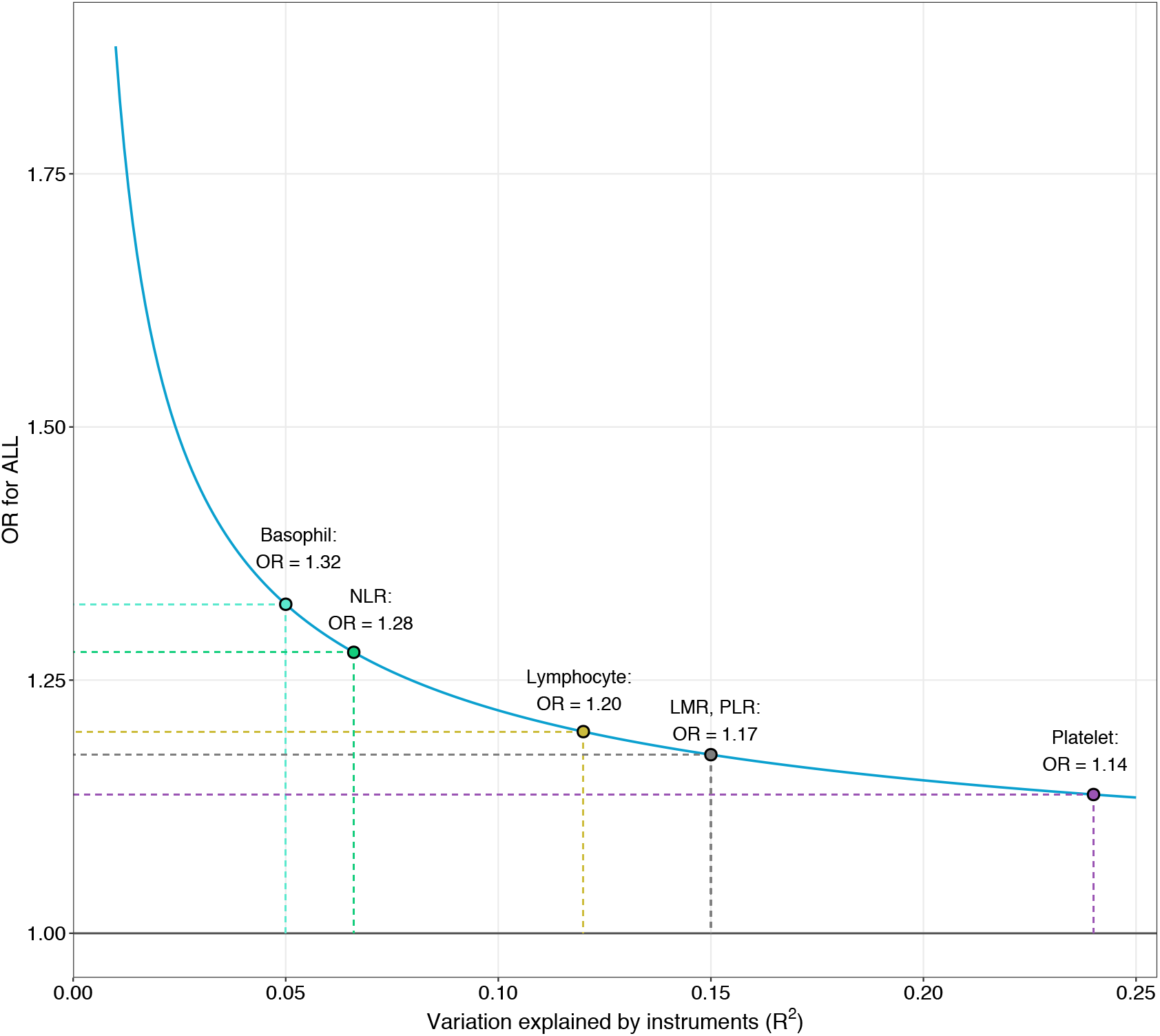
Minimum detectable odds ratio at 80% power for each blood cell trait based on the observed instrument strength and available sample size for the outcome

**Supplementary Figure 6:**
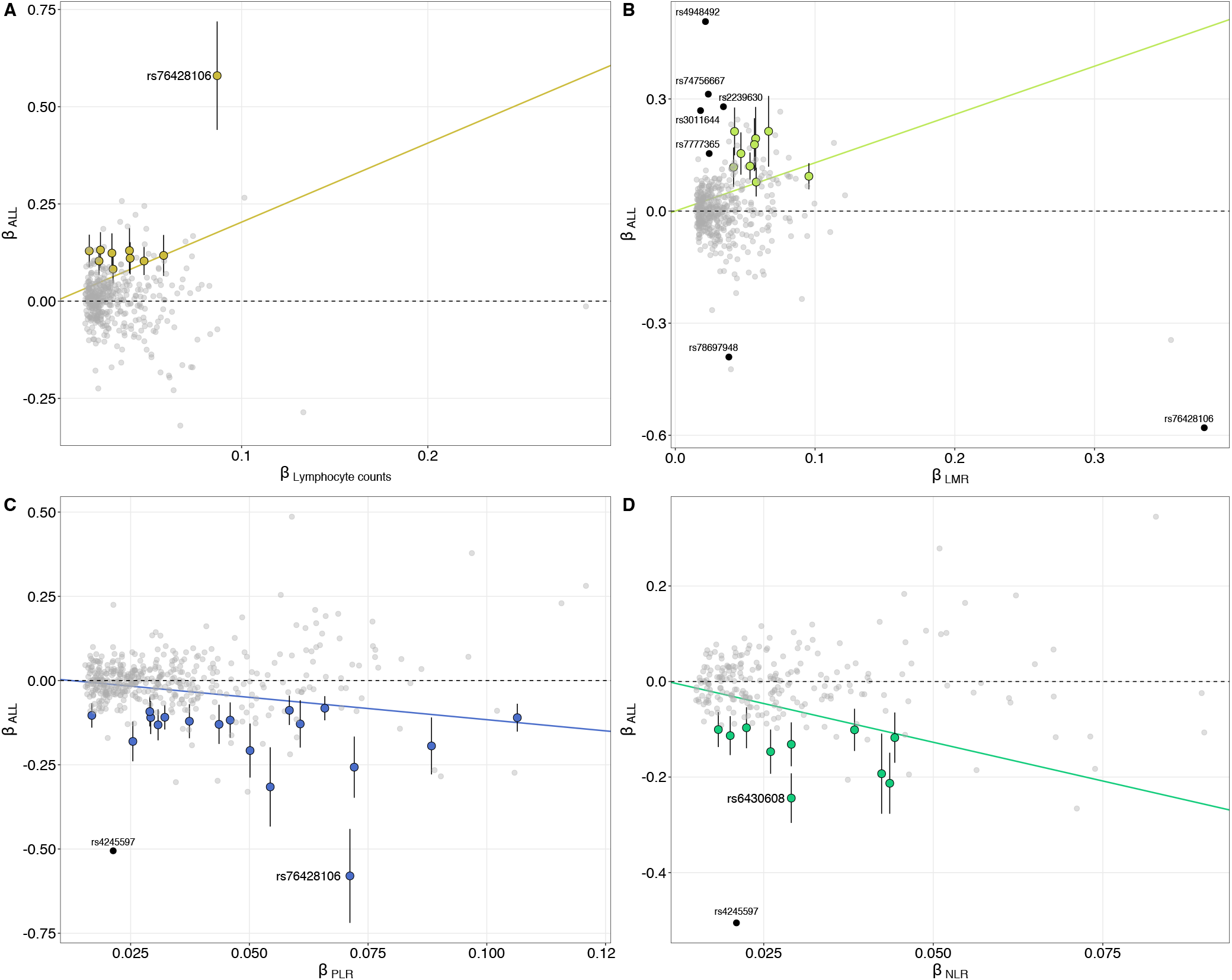
Visualization of MR Clust results identifying subgroups of variants with homogenous causal effects and novel ALL risk variants. The slope of the line corresponds to the mean causal effect within each cluster. Substantive clusters included variants with an assignment probability of greater than 50%. Outliers are denoted by solid black circles. Only variants significantly associated with ALL at the Bonferroni-corrected threshold (*P*_ALL_<5×10^−5^) are labeled.

**Supplementary Table 1:** Array-based heritability (hg) estimates for acute lymphoblastic leukemia (ALL) and blood cell subtypes: lymphocytes, monocytes, neutrophils, basophils, eosinophils, basophils, platelets, lymphocyte to monocyte ratio (LMR), neutrophil to lymphocyte ratio (NLR), and platelet to lymphocyte ratio (PLR) estimated with LD score regression using UKB LD scores from European ancestry participants as the reference panel (7,166,343 variants).

| Phenotype | | $h_g$ | (SE) |
| --- | --- | --- | --- |
| White blood cells |  | 0.141 | (0.009) |
| Lymphocytes |  | 0.165 | (0.014) |
| Monocytes |  | 0.156 | (0.017) |
| Neutrophils |  | 0.122 | (0.011) |
| Eosinophils |  | 0.128 | (0.012) |
| Basophils |  | 0.031 | (0.004) |
| Platelets |  | 0.218 | (0.017) |
| NLR |  | 0.095 | (0.006) |
| LMR |  | 0.117 | (0.011) |
| PLR |  | 0.161 | (0.014) |
| ALL | Lifetime risk: 0.15% | 0.181 | (0.013) |
|  | Lifetime risk: 0.10% | 0.169 | (0.012) |

Note: for ALL heritability was transformed to the liability scale using SEER estimates of lifetime for non-Hispanic whites (0.15%), with a sensitivity analysis using a lower lifetime risk estimate.

**Supplementary Table 2:**
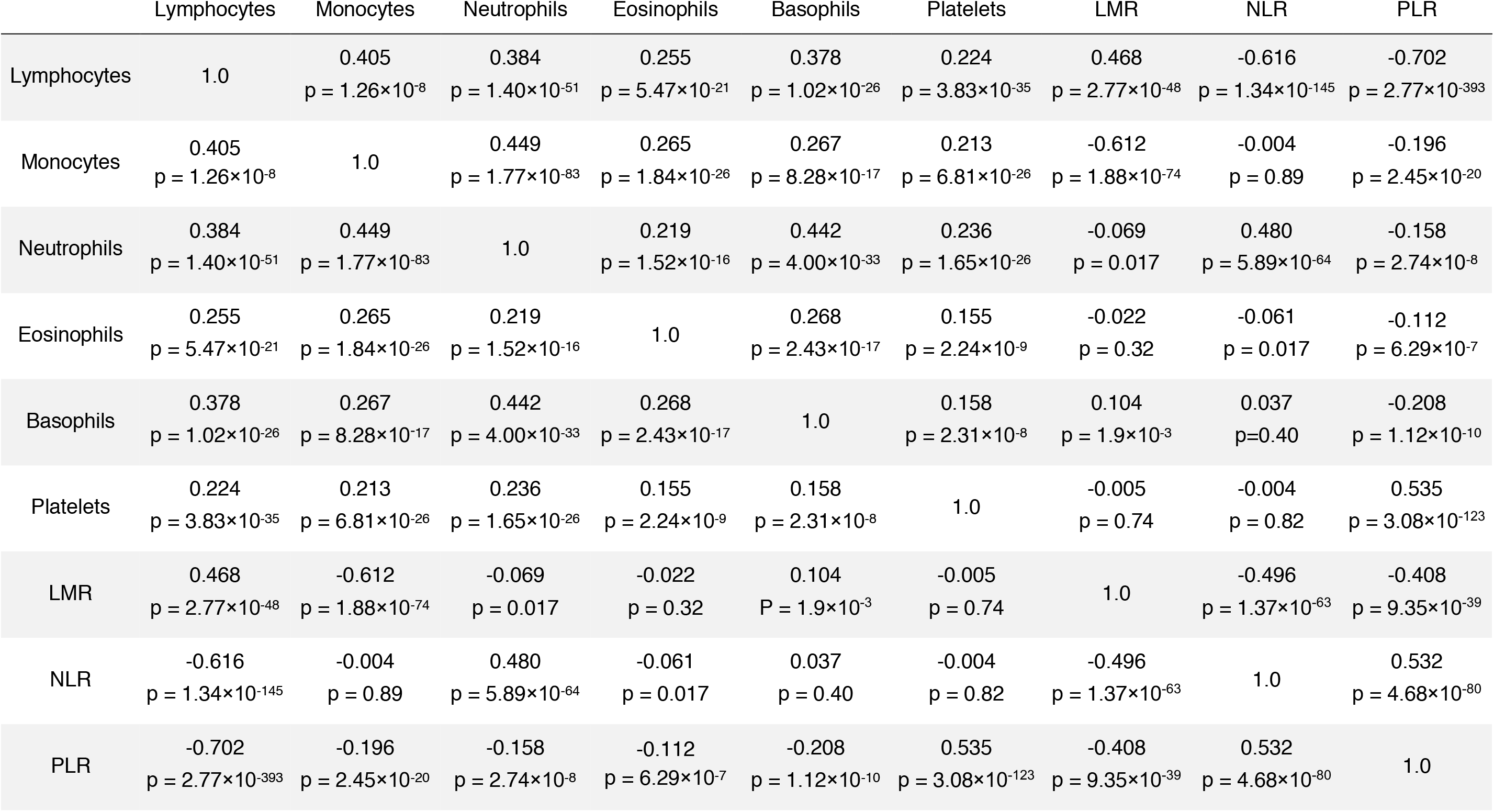
Genetic correlation (*r*_g_) matrix for blood cell subtypes: lymphocytes, monocytes, neutrophils, basophils, eosinophils, basophils, platelets, lymphocyte to monocyte ratio (LMR), neutrophil to lymphocyte ratio (NLR), and platelet to lymphocyte ratio (PLR) estimated using LD score regression. Correlations with p<1.4×10^−3^ were considered statistically significant after Bonferroni correction for 36 pairs tested.

|  | Lymphocytes | Monocytes | Neutrophils | Eosinophils | Basophils | Platelets | LMR | NLR | PLR |
| --- | --- | --- | --- | --- | --- | --- | --- | --- | --- |
| Lymphocytes | 1.0 | 0.405<br>$p = 1.26 \times 10^{-8}$ | 0.384<br>$p = 1.40 \times 10^{-51}$ | 0.255<br>$p = 5.47 \times 10^{-21}$ | 0.378<br>$p = 1.02 \times 10^{-26}$ | 0.224<br>$p = 3.83 \times 10^{-35}$ | 0.468<br>$p = 2.77 \times 10^{-48}$ | -0.616<br>$p = 1.34 \times 10^{-145}$ | -0.702<br>$p = 2.77 \times 10^{-393}$ |
| Monocytes | 0.405<br>$p = 1.26 \times 10^{-8}$ | 1.0 | 0.449<br>$p = 1.77 \times 10^{-83}$ | 0.265<br>$p = 1.84 \times 10^{-26}$ | 0.267<br>$p = 8.28 \times 10^{-17}$ | 0.213<br>$p = 6.81 \times 10^{-26}$ | -0.612<br>$p = 1.88 \times 10^{-74}$ | -0.004<br>$p = 0.89$ | -0.196<br>$p = 2.45 \times 10^{-20}$ |
| Neutrophils | 0.384<br>$p = 1.40 \times 10^{-51}$ | 0.449<br>$p = 1.77 \times 10^{-83}$ | 1.0 | 0.219<br>$p = 1.52 \times 10^{-16}$ | 0.442<br>$p = 4.00 \times 10^{-33}$ | 0.236<br>$p = 1.65 \times 10^{-26}$ | -0.069<br>$p = 0.017$ | 0.480<br>$p = 5.89 \times 10^{-64}$ | -0.158<br>$p = 2.74 \times 10^{-8}$ |
| Eosinophils | 0.255<br>$p = 5.47 \times 10^{-21}$ | 0.265<br>$p = 1.84 \times 10^{-26}$ | 0.219<br>$p = 1.52 \times 10^{-16}$ | 1.0 | 0.268<br>$p = 2.43 \times 10^{-17}$ | 0.155<br>$p = 2.24 \times 10^{-9}$ | -0.022<br>$p = 0.32$ | -0.061<br>$p = 0.017$ | -0.112<br>$p = 6.29 \times 10^{-7}$ |
| Basophils | 0.378<br>$p = 1.02 \times 10^{-26}$ | 0.267<br>$p = 8.28 \times 10^{-17}$ | 0.442<br>$p = 4.00 \times 10^{-33}$ | 0.268<br>$p = 2.43 \times 10^{-17}$ | 1.0 | 0.158<br>$p = 2.31 \times 10^{-8}$ | 0.104<br>$p = 1.9 \times 10^{-3}$ | 0.037<br>$p = 0.40$ | -0.208<br>$p = 1.12 \times 10^{-10}$ |
| Platelets | 0.224<br>$p = 3.83 \times 10^{-35}$ | 0.213<br>$p = 6.81 \times 10^{-26}$ | 0.236<br>$p = 1.65 \times 10^{-26}$ | 0.155<br>$p = 2.24 \times 10^{-9}$ | 0.158<br>$p = 2.31 \times 10^{-8}$ | 1.0 | -0.005<br>$p = 0.74$ | -0.004<br>$p = 0.82$ | 0.535<br>$p = 3.08 \times 10^{-123}$ |
| LMR | 0.468<br>$p = 2.77 \times 10^{-48}$ | -0.612<br>$p = 1.88 \times 10^{-74}$ | -0.069<br>$p = 0.017$ | -0.022<br>$p = 0.32$ | 0.104<br>$p = 1.9 \times 10^{-3}$ | -0.005<br>$p = 0.74$ | 1.0 | -0.496<br>$p = 1.37 \times 10^{-63}$ | -0.408<br>$p = 9.35 \times 10^{-39}$ |
| NLR | -0.616<br>$p = 1.34 \times 10^{-145}$ | -0.004<br>$p = 0.89$ | 0.480<br>$p = 5.89 \times 10^{-64}$ | -0.061<br>$p = 0.017$ | 0.037<br>$p = 0.40$ | -0.004<br>$p = 0.82$ | -0.496<br>$p = 1.37 \times 10^{-63}$ | 1.0 | 0.532<br>$p = 4.68 \times 10^{-80}$ |
| PLR | -0.702<br>$p = 2.77 \times 10^{-393}$ | -0.196<br>$p = 2.45 \times 10^{-20}$ | -0.158<br>$p = 2.74 \times 10^{-8}$ | -0.112<br>$p = 6.29 \times 10^{-7}$ | -0.208<br>$p = 1.12 \times 10^{-10}$ | 0.535<br>$p = 3.08 \times 10^{-123}$ | -0.408<br>$p = 9.35 \times 10^{-39}$ | 0.532<br>$p = 4.68 \times 10^{-80}$ | 1.0 |

**Supplementary Table 3:** Overview of the properties of genetic instruments for blood cell traits. Proportion of trait variation explained was estimated in the independent UK Biobank replication sample of over 100,000 individuals. Instruments applied includes available variants and proxies (LD *r*^2^>0.95) in the outcome GWAS meta-analysis of acute lymphoblastic leukemia.

| Trait | Instruments Available |  | Instruments Applied |  |  |
| --- | --- | --- | --- | --- | --- |
|  | N | Variation (%) | N | Variation (%) | Mean F |
| Lymphocytes | 429 | 11.9 | 406 | 10.7 | 71.1 |
| Monocytes | 505 | 16.9 | 477 | 16.3 | 85.6 |
| Neutrophils | 313 | 9.4 | 301 | 8.7 | 69.4 |
| Eosinophils | 387 | 11.1 | 363 | 10.4 | 71.8 |
| Basophils | 157 | 5.1 | 144 | 4.5 | 64.6 |
| Platelets | 692 | 24.4 | 661 | 23.4 | 85.8 |
| NLR | 266 | 6.6 | 246 | 6.0 | 63.8 |
| LMR | 464 | 15.0 | 432 | 14.4 | 83.8 |
| PLR | 489 | 15.1 | 462 | 14.0 | 76.5 |

**Supplementary Table 4:** Summary of expression quantitative trait loci (eQTL) identified among genetic instruments for blood cell traits.

| Cell Group | eQTL | | eQTL ($P < 5 \times 10^{-8}$) | | | Cell Type | eQTL | | eQTL ($P < 5 \times 10^{-8}$) | | | Study and Sample Size | |
| --- | --- | --- | --- | --- | --- | --- | --- | --- | --- | --- | --- | --- | --- |
|  | N<sub>SNP</sub> | (%) | N<sub>SNP</sub> | % | Genes |  | N<sub>SNP</sub> | % | N<sub>SNP</sub> | % | Genes |  |  |
| Whole Blood | 2405 | (80.2) | 2253 | (75.1) | 2981 | Whole Blood | 2405 | (80.2) | 2253 | (75.1) | 2981 | Vosa bioRxiv: 447367 | 31684 |
| Monocytes (any) | 810 | (27.0) | 514 | (17.1) | 431 | Monocyte | 559 | (18.6) | 370 | (12.3) | 328 | Chen PMID: 27863251 | 197 |
|  |  |  |  |  |  | Monocyte (CD14 classical) | 376 | (12.5) | 231 | (7.7) | 148 | Momozawa PMID: 29930244 | 322 |
|  |  |  |  |  |  | Monocyte (CD14 classical) | 228 | (7.6) | 83 | (2.8) | 68 | Schmiedel PMID: 30449622 | 91 |
|  |  |  |  |  |  | Monocyte (non-classical) | 206 | (6.9) | 64 | (2.1) | 58 | Schmiedel PMID: 30449622 | 91 |
| Neutrophils (any) | 633 | (21.1) | 419 | (14) | 341 | Neutrophil | 458 | (15.3) | 289 | (9.6) | 274 | Chen PMID: 27863251 | 197 |
|  |  |  |  |  |  | Neutrophil (CD15) | 391 | (13) | 253 | (8.4) | 168 | Momozawa PMID: 29930244 | 322 |
| T-cells (any) | 708 | (23.6) | 362 | (12.1) | 327 | T-cell | 430 | (14.3) | 277 | (9.2) | 233 | Chen PMID: 27863251 | 197 |
|  |  |  |  |  |  | T CD8 (naïve) | 221 | (7.4) | 87 | (2.9) | 84 | Schmiedel PMID: 30449622 | 91 |
|  |  |  |  |  |  | T CD4 TH17 | 242 | (8.1) | 78 | (2.6) | 75 | Schmiedel PMID: 30449622 | 91 |
|  |  |  |  |  |  | T_CD4 (memory TREG) | 230 | (7.7) | 79 | (2.6) | 73 | Schmiedel PMID: 30449622 | 91 |
|  |  |  |  |  |  | T_CD4 (naïve TREG) | 228 | (7.6) | 77 | (2.6) | 79 | Schmiedel PMID: 30449622 | 91 |
|  |  |  |  |  |  | T_CD4_TH2 | 222 | (7.4) | 74 | (2.5) | 69 | Schmiedel PMID: 30449622 | 91 |
|  |  |  |  |  |  | T_CD4 (naïve) | 223 | (7.4) | 69 | (2.3) | 71 | Schmiedel PMID: 30449622 | 91 |
|  |  |  |  |  |  | T_CD4_TFH | 216 | (7.2) | 66 | (2.2) | 65 | Schmiedel PMID: 30449622 | 91 |
|  |  |  |  |  |  | T_CD4_TH1_17 | 207 | (6.9) | 64 | (2.1) | 61 | Schmiedel PMID: 30449622 | 91 |
|  |  |  |  |  |  | T_CD4_TH1 | 182 | (6.1) | 51 | (1.7) | 45 | Schmiedel PMID: 30449622 | 91 |
|  |  |  |  |  |  | T_CD8 (naïve activated) | 148 | (4.9) | 48 | (1.6) | 39 | Schmiedel PMID: 30449622 | 91 |
|  |  |  |  |  |  | T_CD4 (naïve activated) | 138 | (4.6) | 45 | (1.5) | 42 | Schmiedel PMID: 30449622 | 91 |
| B-cells (any) | 341 | (11.4) | 166 | (5.5) | 119 | B-cell (CD19) | 233 | (7.8) | 125 | (4.2) | 69 | Momozawa PMID: 29930244 | 322 |
|  |  |  |  |  |  | B-cell (naïve) | 185 | (6.2) | 73 | (2.4) | 62 | Schmiedel PMID: 30449622 | 91 |
| Platelets | 119 | (4.0) | 72 | (2.4) | 51 | Platelets | 119 | (4.0) | 72 | (2.4) | 51 | Momozawa PMID: 29930244 | 322 |
| NK cells | 154 | (5.1) | 48 | (1.6) | 44 | NK cells | 154 | (5.1) | 48 | (1.6) | 44 | Schmiedel PMID: 30449622 | 91 |

**Supplementary Table 5:** Odds ratios (OR) and 95% confidence intervals (CI) for the effect of increasing blood cell counts or cell type ratios on the risk of acute lymphoblastic leukemia (ALL), estimated using Mendelian Randomization (MR).

| Trait | MR Estimator | N<sub>SNP</sub> | Association Results |  |  |
| --- | --- | --- | --- | --- | --- |
|  |  |  | OR | (95% CI) | P |
| Basophils | IVW (multiplicative random effects) | 144 | 0.95 | (0.59 – 1.53) | 0.83 |
|  | Maximum likelihood |  | 0.95 | (0.66 – 1.36) | 0.77 |
|  | Weighted median |  | 1.31 | (0.71 – 2.39) | 0.39 |
|  | PRESSO (corrected) |  | 0.94 | (0.62 – 1.43) | 0.78 |
|  | RAPS shrinkage |  | 0.97 | (0.67 – 1.41) | 0.86 |
| Eosinophils | IVW (multiplicative random effects) | 363 | 1.01 | (0.84 – 1.22) | 0.91 |
|  | Maximum likelihood |  | 1.01 | (0.86 – 1.18) | 0.89 |
|  | Weighted median |  | 0.84 | (0.64 – 1.09) | 0.19 |
|  | PRESSO (corrected) |  | 1.00 | (0.84 – 1.21) | 0.97 |
|  | RAPS shrinkage |  | 0.95 | (0.81 – 1.12) | 0.55 |
| Lymphocytes | IVW (multiplicative random effects) | 406 | 1.15 | (0.99 – 1.34) | 0.061 |
|  | Maximum likelihood |  | 1.16 | (1.01 – 1.33) | 0.035 |
|  | Weighted median |  | 0.96 | (0.71 – 1.29) | 0.78 |
|  | PRESSO (corrected) |  | 1.14 | (0.98 – 1.32) | 0.087 |
|  | RAPS shrinkage |  | 1.16 | (1.01 – 1.34) | 0.033 |
| Monocytes | IVW (multiplicative random effects) | 477 | 1.01 | (0.98 – 1.05) | 0.49 |
|  | Maximum likelihood |  | 1.01 | (0.98 – 1.04) | 0.40 |
|  | Weighted median |  | 1.02 | (0.99 – 1.05) | 0.25 |
|  | PRESSO (corrected) |  | 1.01 | (0.98 – 1.04) | 0.52 |
|  | RAPS shrinkage |  | 1.01 | (0.98 – 1.04) | 0.39 |
| Neutrophils | IVW (multiplicative random effects) | 301 | 1.04 | (0.85 – 1.27) | 0.70 |
|  | Maximum likelihood |  | 1.04 | (0.87 – 1.25) | 0.67 |
|  | Weighted median |  | 1.08 | (0.80 – 1.46) | 0.62 |
|  | PRESSO (corrected) |  | 1.00 | (0.83 – 1.22) | 0.97 |
|  | RAPS shrinkage |  | 1.01 | (0.84 – 1.22) | 0.93 |
| Platelets | IVW (multiplicative random effects) | 661 | 1.02 | (0.97 – 1.06) | 0.51 |
|  | Maximum likelihood |  | 1.02 | (0.97 – 1.06) | 0.47 |
|  | Weighted median |  | 1.02 | (0.97 – 1.08) | 0.36 |
|  | PRESSO (corrected) |  | 1.02 | (0.97 – 1.07) | 0.40 |
|  | RAPS shrinkage |  | 1.02 | (0.97 – 1.06) | 0.49 |
| LMR | IVW (multiplicative random effects) | 432 | 1.22 | (1.00 – 1.50) | 0.052 |
|  | Maximum likelihood |  | 1.23 | (1.07 – 1.41) | 4.5×10<sup>-3</sup> |
|  | Weighted median |  | 1.02 | (0.80 – 1.31) | 0.86 |
|  | PRESSO (corrected) |  | 1.18 | (1.01 – 1.38) | 0.037 |
|  | RAPS shrinkage |  | 1.14 | (0.99 – 1.32) | 0.070 |
| NLR | IVW (multiplicative random effects) |  | 0.67 | (0.49 – 0.92) | 0.012 |
| | Maximum likelihood | | 0.67 | (0.54 – 0.83) | $3.1 \times 10^{-4}$ |
|  | Weighted median | 246 | 0.66 | (0.47 – 0.93) | 0.017 |
|  | PRESSO (corrected) |  | 0.77 | (0.60 – 0.98) | 0.036 |
| | RAPS shrinkage | | 0.73 | (0.58 – 0.91) | $5.6 \times 10^{-3}$ |
| PLR | IVW (multiplicative random effects) |  | 0.80 | (0.67 – 0.96) | 0.018 |
| | Maximum likelihood | | 0.80 | (0.70 – 0.92) | $2.0 \times 10^{-3}$ |
|  | Weighted median | 462 | 0.85 | (0.67 – 1.08) | 0.18 |
|  | PRESSO (corrected) |  | 0.82 | (0.70 – 0.96) | 0.014 |
|  | RAPS shrinkage |  | 0.85 | (0.73 – 0.98) | 0.025 |

**Supplementary Table 6:** Summary of diagnostic tests carried out for Mendelian Randomization analyses

| Trait | Heterogeneity<sup>1</sup> | | | MR Egger<sup>2</sup> | | MR PRESSO | | $I^2_{GX}$<sup>5</sup> | $P_{Steiger}$<sup>6</sup> |
| --- | --- | --- | --- | --- | --- | --- | --- | --- | --- |
| | Q | DF | $P_{Q-value}$ | $\beta_0$ | $P_{Egger}$ | $P_{Global}$<sup>3</sup> | $P_{Dist}$<sup>4</sup> | | |
| Basophils | 262.3 | 143 | $4.7 \times 10^{-9}$ | -0.0099 | 0.40 | $<6.7 \times 10^{-5}$ | 0.98 | 0.985 | $1.8 \times 10^{-14}$ |
| Eosinophils | 509.9 | 360 | $3.1 \times 10^{-7}$ | 0.0028 | 0.69 | $<6.7 \times 10^{-5}$ | 0.04 | 0.986 | $1.6 \times 10^{-75}$ |
| Lymphocytes | 512.1 | 398 | $9.4 \times 10^{-5}$ | 0.0082 | 0.09 | $<6.7 \times 10^{-5}$ | 0.88 | 0.986 | $5.0 \times 10^{-135}$ |
| Monocytes | 708.8 | 472 | $9.0 \times 10^{-12}$ | -0.0040 | 0.15 | $2.7 \times 10^{-4}$ | 0.69 | 0.988 | $1.8 \times 10^{-269}$ |
| Neutrophils | 368.3 | 299 | $3.8 \times 10^{-3}$ | -0.0098 | 0.16 | $3.7 \times 10^{-3}$ | 0.03 | 0.986 | $3.8 \times 10^{-120}$ |
| Platelets | 764.5 | 654 | $1.8 \times 10^{-3}$ | -0.0002 | 0.93 | $3.1 \times 10^{-3}$ | 0.75 | 0.988 | $<1 \times 10^{-300}$ |
| LMR | 899.0 | 428 | $1.2 \times 10^{-35}$ | 0.0097 | 0.15 | $<6.7 \times 10^{-5}$ | 0.63 | 0.988 | $5.4 \times 10^{-98}$ |
| NLR | 524.1 | 242 | $6.8 \times 10^{-23}$ | -0.0103 | 0.39 | $<6.7 \times 10^{-5}$ | 0.15 | 0.984 | $2.1 \times 10^{-10}$ |
| PLR | 798.9 | 456 | $4.1 \times 10^{-21}$ | -0.0071 | 0.30 | $<6.7 \times 10^{-5}$ | 0.83 | 0.987 | $1.2 \times 10^{-109}$ |

<sup>1</sup> Cochran's Q based on modified second order weights, p-values<0.05 indicate statistically significant heterogeneity
<sup>2</sup> Presence of statistically significant directional pleiotropy is indicated by non-zero intercept values with p<0.05
<sup>3</sup> Estimated based on 15000 replicates; p-values<0.05 indicate statistically significant pleiotropy
<sup>4</sup> Estimated based on 15000 replicates; p-values<0.05 indicate statistically significant distortion in causal effect estimates
<sup>5</sup> $I^2_{GX}$ <0.90 indicates weak instrument bias due to violations of the no exposure measurement error assumption
<sup>6</sup> Steiger directionality test for orienting the causal effect as exposure → outcome

**Supplementary Table 7:**
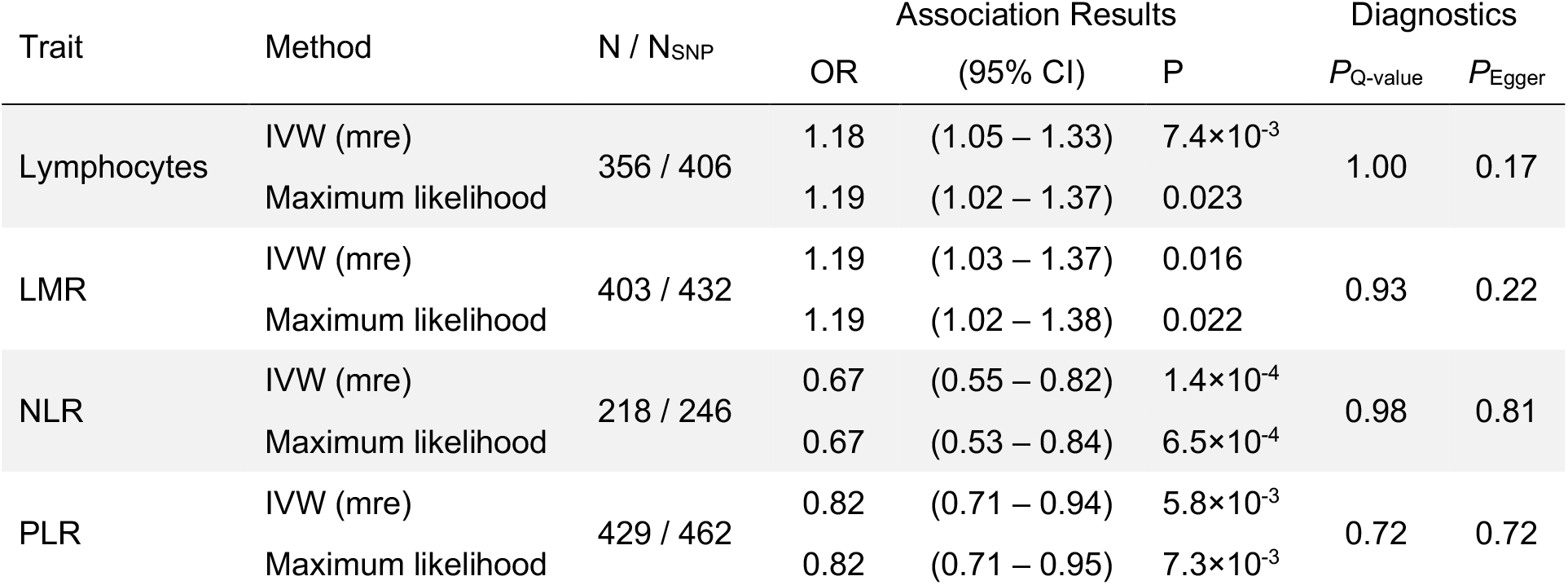
Odds ratios (OR) and 95% for acute lymphoblastic leukemia (ALL), estimated using Mendelian Randomization (MR) for selected traits after filtering out instruments that significantly contributed to heterogeneity.

**Supplementary Table 8:**
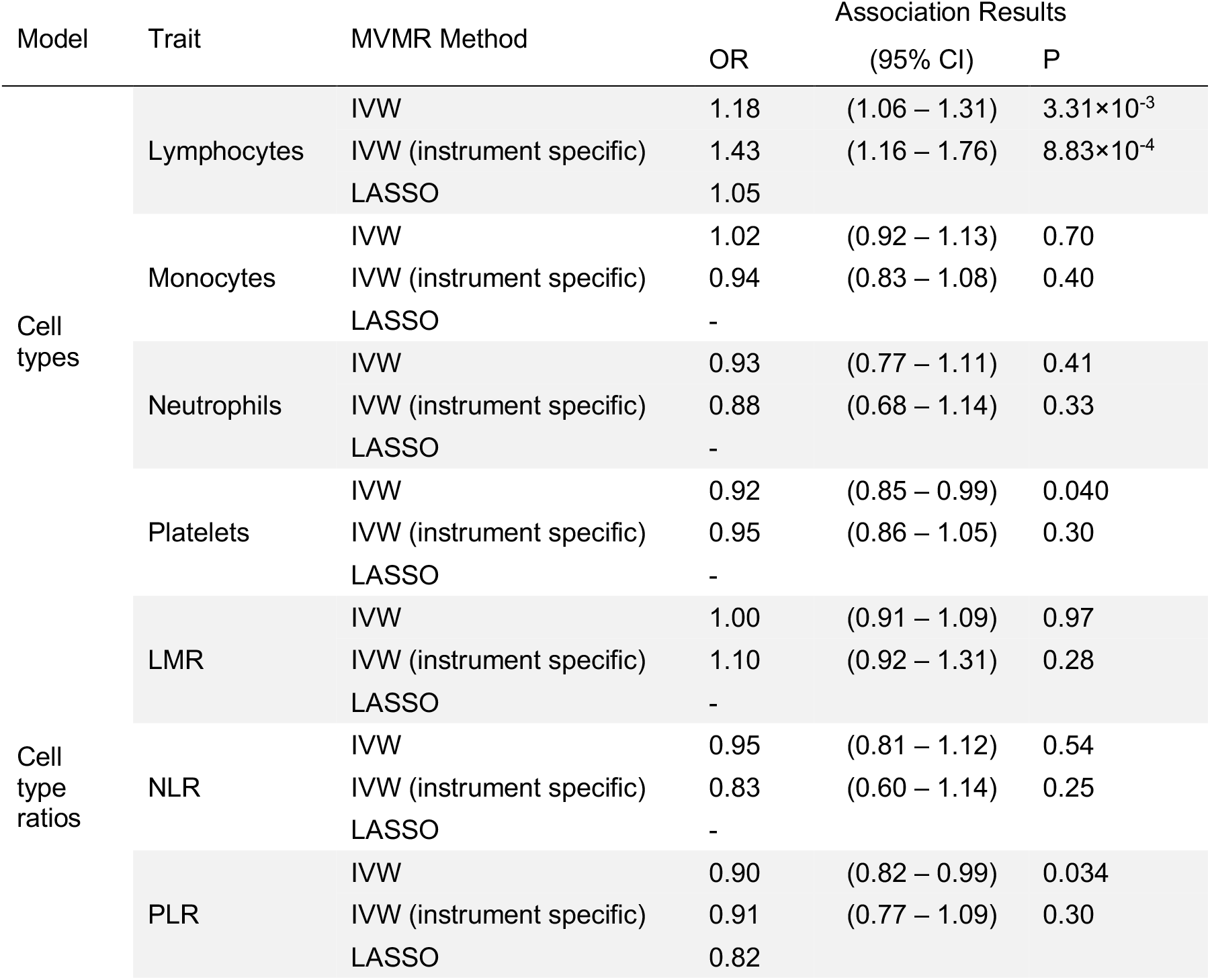
Odds ratios (OR) and 95% for acute lymphoblastic leukemia (ALL), estimated using multivariable Mendelian Randomization (MVMR) for traits that were individually associated with ALL or were used to derive relevant ratio phenotypes.

**Supplementary Table 9:**
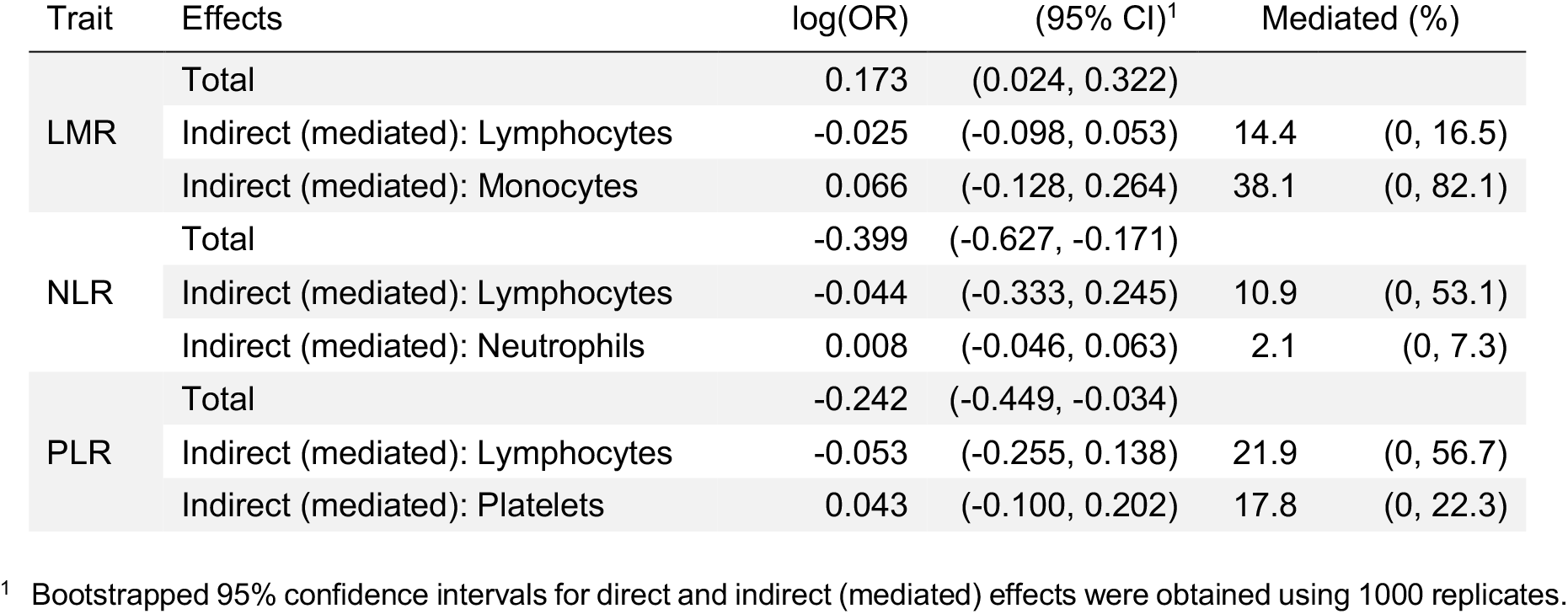
Mediation analyses that decompose the total effect of blood cell ratios on ALL risk into effects mediated by their component cell types. Total effects are based on Mendelian randomization results after filtering out instruments that significantly contributed to heterogeneity, as seen in Supplementary Table 6.

**Supplementary Table 10:**
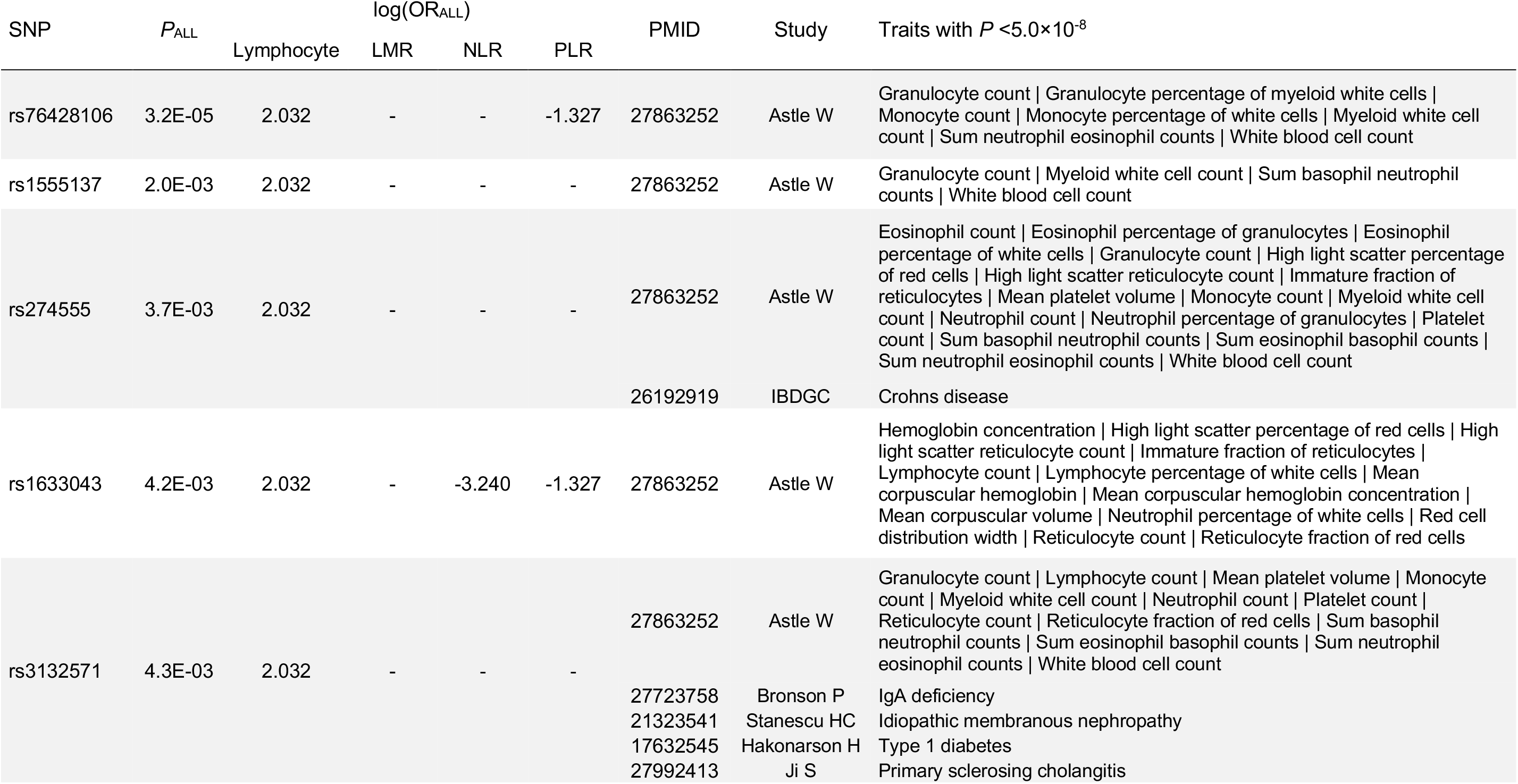
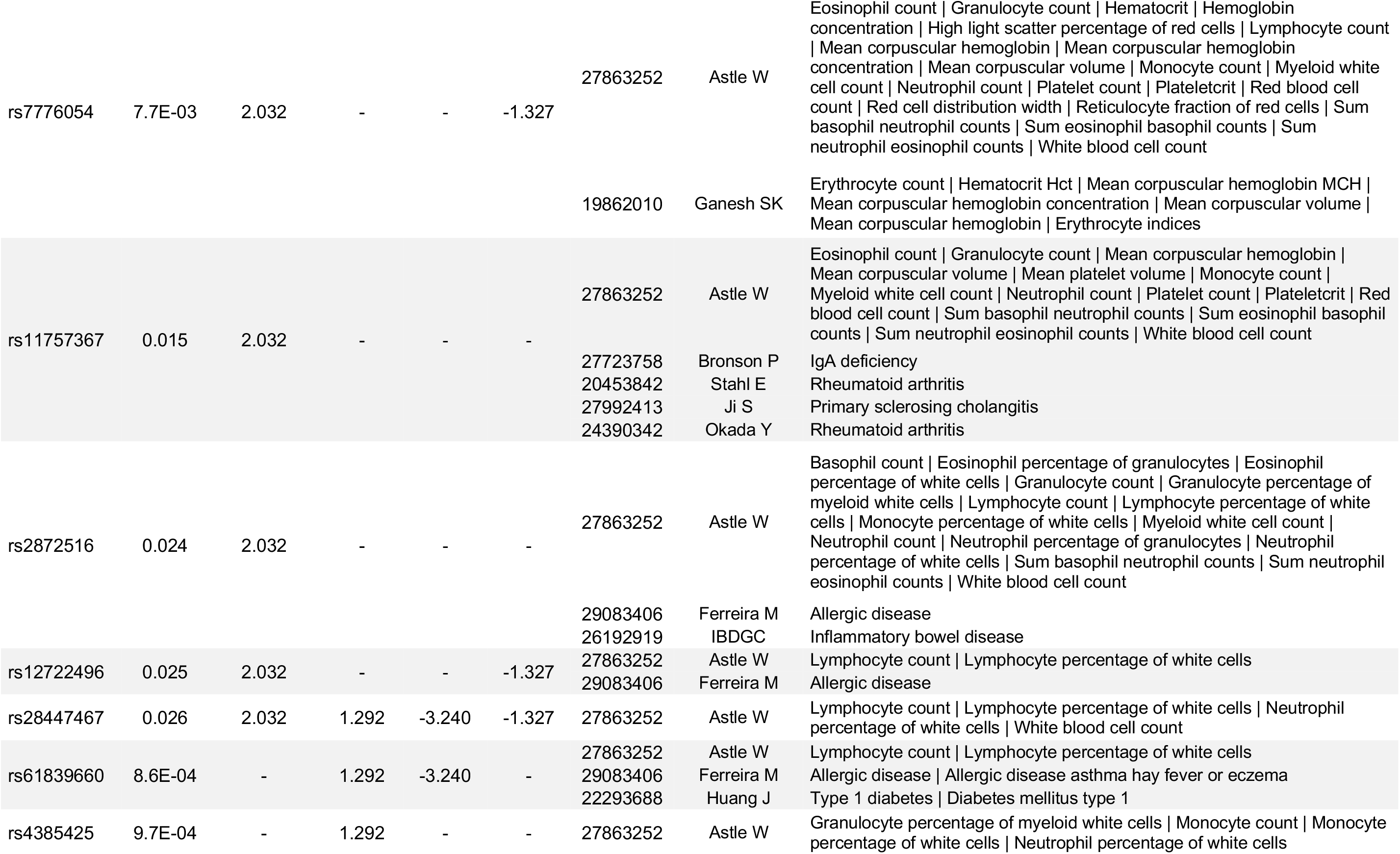
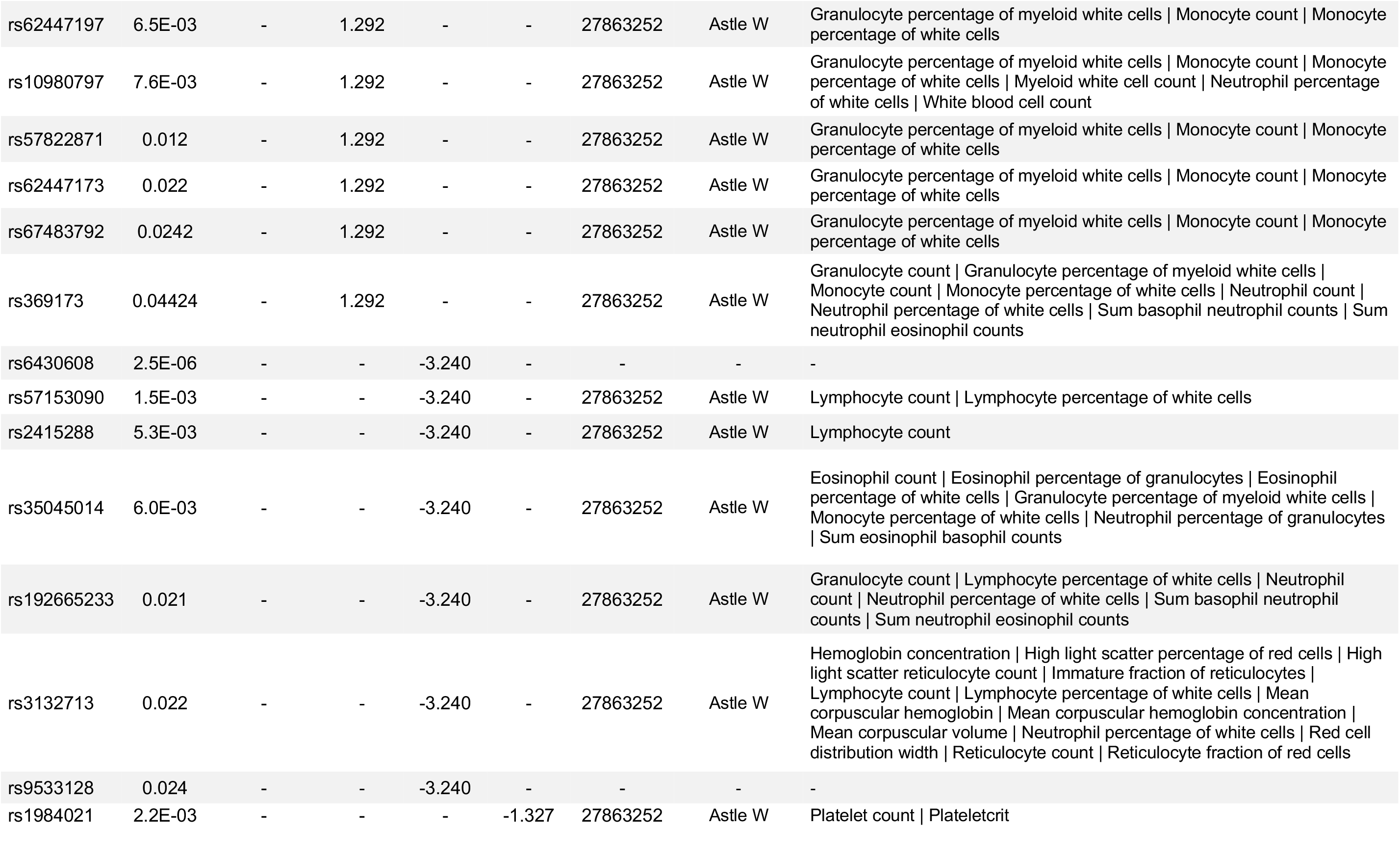
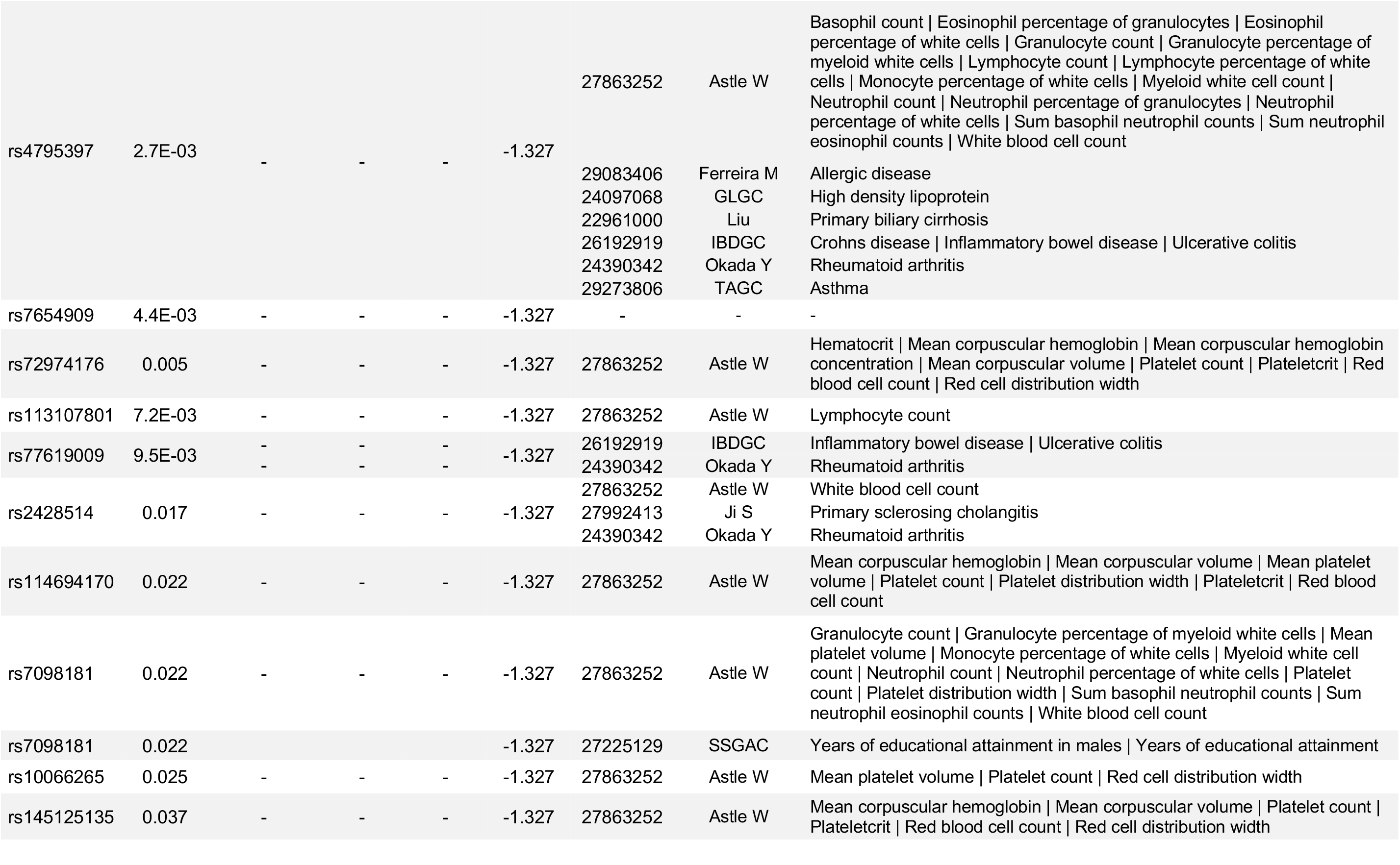
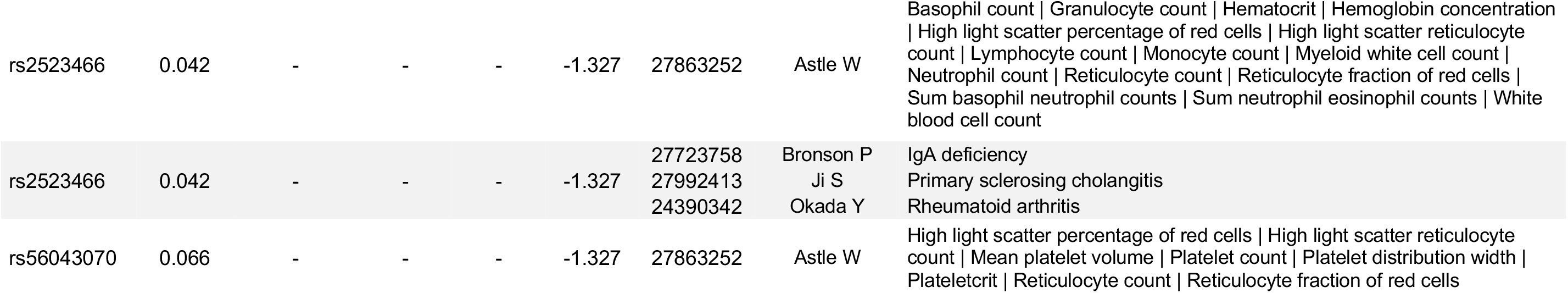
Annotation of blood cell trait instruments assigned to substantive clusters using the MR Clust algorithm with an assignment probability of greater than 50%. For each blood cell trait, the substantive cluster mean corresponds to the causal effect on ALL, log odds ratio (OR) per 1 unit increase, indicated by the variants comprising that cluster. Published associations with other phenotypes were obtained by querying the PhenoScanner database.

**Supplementary Table 11:** Mediation analyses that decompose the total effect of selected ALL risk variants into direct and indirect effects, mediated via regulation of blood cell profiles. Mediator-outcome effects were obtained from Mendelian randomization analyses excluding outliers (as seen in Supplementary Table 7).

|  |  | OR<sub>ALL</sub> | P<sub>ALL</sub> | β<sub>ALL</sub> | (95% CI)<sup>1</sup> | Mediated (%) |  |
| --- | --- | --- | --- | --- | --- | --- | --- |
| Mediator: PLR |  | 0.82 | 5.8×10<sup>-3</sup> | -0.200 |  |  |  |
| rs4245597<br>(ARID5B, 10q21.2) | Total | 0.60 | 5.3×10<sup>-46</sup> | -0.505 | (-0.575, -0.436) |  |  |
|  | Direct |  |  | -0.500 | (-0.571, -0.431) |  |  |
|  | Indirect (mediated) |  |  | -0.004 | (-0.007, -0.001) | 0.84 | (0.23, 1.44) |
| rs76428106<br>(FLT3, 13q12.2) | Total | 0.56 | 3.2×10<sup>-5</sup> | -0.580 | (-0.853, -0.307) |  |  |
|  | Direct |  |  | -0.562 | (-0.841, -0.291) |  |  |
|  | Indirect (mediated) |  |  | -0.014 | (-0.024, -0.004) | 2.43 | (0.68, 4.19) |
| Mediator: NLR |  | 0.67 | 1.4×10<sup>-4</sup> | -0.400 |  |  |  |
| rs4245597<br>(ARID5B, 10q21.2) | Total | 0.60 | 5.3×10<sup>-46</sup> | -0.505 | (-0.575, -0.436) |  |  |
|  | Direct |  |  | -0.496 | (-0.567, -0.426) |  |  |
|  | Indirect (mediated) |  |  | -0.008 | (-0.013, -0.004) | 1.65 | (0.79, 2.51) |
| rs6430608<br>(2q22.1) | Total | 0.78 | 2.5×10<sup>-6</sup> | -0.244 | (-0.346, -0.142) |  |  |
|  | Direct |  |  | -0.231 | (-0.334, -0.129) |  |  |
|  | Indirect (mediated) |  |  | -0.012 | (-0.018, -0.006) | 4.72 | (2.26, 7.20) |
| Mediator: LMR |  | 1.19 | 0.016 | 0.173 |  |  |  |
| rs4948492<br>(ARID5B, 10q21.2) | Total | 1.66 | 5.1×10<sup>-46</sup> | 0.507 | (0.437, 0.577) |  |  |
|  | Direct |  |  | 0.503 | (0.433, 0.574) |  |  |
|  | Indirect (mediated) |  |  | 0.004 | (-0.001, 0.008) | 0.75 | (0, 1.63) |
| rs74756667<br>(8q24.2) | Total | 1.37 | 3.3×10<sup>-10</sup> | 0.313 | (0.215, 0.410) |  |  |
|  | Direct |  |  | 0.310 | (0.210, 0.407) |  |  |
|  | Indirect (mediated) |  |  | 0.004 | (0.001, 0.007) | 1.31 | (0.24, 2.38) |
| rs2239630<br>(CEBPE, 14q11.2) | Total | 1.32 | 1.2×10<sup>-13</sup> | 0.278 | (0.206, 0.353) |  |  |
|  | Direct |  |  | 0.275 | (0.199, 0.348) |  |  |
|  | Indirect (mediated) |  |  | 0.006 | (-0.001, 0.013) | 2.16 | (0, 4.71) |
| rs78697948*<br>(IKZF1, 7p12.2) | Total | 0.68 | 1.2×10<sup>-10</sup> | -0.391 | (-0.509, -0.272) |  |  |
|  | Direct |  |  | -0.395 | (-0.516, -0.276) |  |  |
|  | Indirect (mediated) |  |  | 0.007 | (-0.001, 0.015) | 1.72 | (0, 3.75) |
| rs76428106*<br>(FLT3, 13q12.2) | Total | 0.56 | 3.2×10<sup>-5</sup> | -0.580 | (-0.853, -0.307) |  |  |
|  | Direct |  |  | -0.637 | (-0.918, -0.368) |  |  |
|  | Indirect (mediated) |  |  | 0.066 | (0.012, 0.120) | 11.39 | (2.07, 20.7) |
| Mediator: Lymphocytes |  | 1.18 | 7.4×10<sup>-3</sup> | 0.166 | (0.045, 0.288) |  |  |
| rs76428106<br>(FLT3, 13q12.2) | Total | 1.79 | 3.2×10<sup>-5</sup> | 0.580 | (0.307, 0.853) |  |  |
|  | Direct |  |  | 0.569 | (0.290, 0.840) |  |  |
|  | Indirect (mediated) |  |  | 0.015 | (0.004, 0.025) | 2.51 | (0.67, 4.35) |

<sup>1</sup> Bootstrapped 95% confidence intervals for direct and indirect (mediated) effects were obtained using 1000 replicates.
\* Variants had statistically significant effects, and of larger magnitude, on monocyte counts than LMR or lymphocytes

