## Supplementary Note for "Genetic Determinants of Blood Cell Traits Influence Susceptibility to Childhood Acute Lymphoblastic Leukemia"

### Childhood Acute Lymphoblastic Leukemia GWAS Meta-Analysis Methods

### CCRLP/GERA Study Subjects

The California Cancer Records Linkage Project (CCRLP) includes all children born in California during 1982-2009 and diagnosed with acute lymphoblastic leukemia (ALL) at the age of 0-14 years per California Cancer Registry records data[1]. Children born in California during the same time period and not reported to the California Cancer Registry as having any childhood cancer were considered as potential controls. Up to four control subjects were randomly selected for each case from a pool of potential controls and matched to case on year and month of birth, sex, and race/ethnicity. Newborn dried bloodspot samples (DBS) were obtained for childhood ALL cases and an approximately equal number of controls, selected randomly out of the up to four matched controls, from the California Biobank Program. Further details, including on DNA isolations from newborn DBS and SNP array genotyping, have been previously described[1].

Additional California-based controls were included from the Kaiser Resource for Genetic Epidemiology Research on Aging Cohort (GERA)[2]. The GERA cohort was chosen due to the similar use of the Affymetrix Axiom World Array platform. The GERA genotype data were downloaded from dbGaP (Study Accession: phs000788.v1.p2).

Self-reported race/ethnicity was available for all subjects in the CCRLP and GERA studies. We did not attempt to reassign individuals to different race/ethnic groups based on estimated genetic ancestry. Principal components analysis was performed including CCRLP/GERA subjects along with 1000 Genomes Project reference data to identify extreme outlier individuals that clustered apart from other individuals in their self-reported race/ethnicity groups: this did not identify any self-reported non-Latino white subjects that clustered with individuals in non-European ancestry populations in 1000 Genomes data. Thus, following quality control (see below), there were 1,162 cases and 1,229 controls in CCRLP and 56,112 controls in GERA of non-Latino white self-reported race/ethnicity that we designated as European ancestry individuals to be included in the GWAS of childhood ALL.

### COG/WTCCC Study Subjects

The Children's Oncology Group (COG) study genotype data were downloaded from dbGaP (Study Accession: phs000638.v1.p1) and include childhood ALL cases genotyped on the Affymetrix Human SNP Array 6.0 (COG trials AALL0232 and P9904/9905) or the Affymetrix GeneChip Human Mapping 500K Array (COG P9906 and St. Jude Total Therapy XIIIB/XV)[3,4,5]. For controls, genotype data were downloaded from individuals from the Wellcome Trust Case-Control Consortium (WTCCC), also genotyped on the Affymetrix Human SNP Array 6.0[6]. Self-reported race/ethnicity was not available for COG/WTCCC subjects, therefore we performed global ancestry estimations using ADMIXTURE and the 1000 Genomes populations as reference, and removed individuals with < 90% estimated European ancestry from the analysis. This resulted in 1,504 cases in the COG and 2,931 controls in the WTCCC datasets.

**Data Processing and Quality Control**

Quality control (QC) procedures for SNP array genotypes and samples were carried out in each population and dataset in parallel, performed in two stages: pre-imputation and post-imputation. In pre-imputation QC, the sex chromosomes were excluded, and SNPs were filtered out on the basis of call rate (<98%), minor allele frequency (MAF<0.01), genome-wide relatedness (PI_HAT>0.02), genome heterozygosity rate (mean heterozygosity± 6Std), and deviation from Hardy-Weinberg equilibrium in controls ($P_{HWE}<1x10^{-5}$). Samples with call rate < 95% were also removed. To control for potential batch effect and systematic bias between array types, we performed two separate GWASs in CCRLP/GERA. First, restricted to post-QC SNPs in both CCRLP and GERA, we compared non-Latino white CCRLP controls and GERA individuals. Second, in the GERA controls we compared individuals that were genotyped on the Axiom type "A" to those genotyped on the type "O" reagent kit. 20 principal components (PCs) were included as covariates in logistic regression tests. In both comparisons we observed inflation of the test statistics suggesting a subset of SNPs exhibited evidence of batch effect, thus we removed variants with P < 0.01 in each of the comparisons.

Genome-wide imputation was performed in each dataset using the Haplotype Reference Consortium (HRC v r1.1 2016)[7] as a reference in the Michigan Imputation Server. In post-imputation QC, we filtered out variants by imputation quality (INFO score < 0.3), MAF (< 0.01), and allele frequency difference (> 0.1) between CCRLP and WTCCC controls and the non-Finnish European population in the Genome Aggregation Database (gnomAD)[8]. We next performed another GWAS between CCRLP controls and GERA individuals, and removed variants with $P < 1x10^{-5}$.

**ALL GWAS Meta-Analysis**

We merged the CCRLP with the GERA dataset and the COG with the WTCCC dataset to perform two separate GWAS of ALL. In each case-control study, we used SNPTEST[9] (v2.5.2) to test the association between imputed genotype dosage at each SNP and case-control status in logistic regression, after adjusting for the top 20 PCs. The results from the CCRLP/GERA and COG/WTCCC cohort were combined via the fixed-effect meta-analysis with variance weighting using METAL[10].
